# A novel Bayesian fine-mapping model using a continuous global-local shrinkage prior with applications in prostate cancer analysis

**DOI:** 10.1101/2023.08.04.23293456

**Authors:** Xiang Li, Pak Chung Sham, Yan Dora Zhang

## Abstract

The aim of fine-mapping is to identify genetic variants causally contributing to complex traits or diseases. Existing fine-mapping methods employ discrete Bayesian mixture priors and depend on a pre-specified maximum number of causal variants which may lead to sub-optimal solutions. In this work, we propose a novel fine-mapping method called h2-D2, utilizing a continuous global-local shrinkage prior. We also present an approach to define credible sets of causal variants in continuous prior settings. Simulation studies demonstrate that h2-D2 outperforms the state-of-art fine-mapping methods such as SuSiE and FINEMAP in accurately identifying causal variants and estimating their effect sizes. We further applied h2-D2 to prostate cancer analysis and discovered some previously unknown causal variants. In addition, we inferred 385 target genes associated with the detected causal variants and several pathways that were significantly over-represented by these genes, shedding light on their potential roles in prostate cancer development and progression.

## Introduction

Genome-wide association studies (GWAS) have discovered numerous genetic variants associated with a wide range of complex traits and diseases [1]. However, pinpointing the specific variants that have causal effects on the traits is challenging due to the presence of high linkage disequilibrium (LD) among single nucleotide polymorphisms (SNPs) and their small effect sizes [2–4]. The goal of statistical fine-mapping is to identify the causal variants that have nonzero effects on the trait, which is essentially a statistical problem known as “variable selection”. Since it is difficult to distinguish a causal variant from other variants highly correlated with it without extra information, penalized regression methods sometimes fail to select the true causal variants [5]. On the other hand, Bayesian methods are more appropriate for fine-mapping by providing posterior “credible sets” (CSs) [4]. A level 1 −*α* CS is defined as a set of variants that contains at least one causal variant with posterior probability no less than 1 −*α* [6, 7]. A CS may contain multiple highly correlated candidate causal variants for further functional validation.

To date, many Bayesian fine-mapping methods have been developed, including CAVIAR [2], CAVIARBF [5], PAINTOR [3], JAM [8], DAP [9], FINEMAP [10, 11], and SuSiE [12]. All these methods are based on discrete mixture priors, specifying a prior probability for each variant being causal. Suppose there are *M* SNPs in the region of interest, the number of possible models is 2^*M*^ . To reduce computational cost, these methods need to set limit on the maximum number of causal variants. However, mis-specifying the number may lead to decrease in performance[12]. In addition, existing methods rely on exhaustive search, shotgun stochastic search, or stepwise selection to explore the space of causal configurations, which can be time-consuming or lead to poor suboptimal solutions [6, 12].

In Bayesian analysis, there is another class of shrinkage priors termed “continuous global-local shrinkage priors”. Existing continuous priors have been shown to be efficient variable selection tools [13–20] and have been successfully applied in genetic studies, including polygenic risk prediction [21]. However, continuous shrinkage priors are hardly used in fine-mapping. One shortcoming of continuous priors is that they require additional procedures in order to perform variable selection, as the posterior mean of regression coefficients is not sparse almost surely. Existing approaches include hard thresholding methods [15, 22], penalized credible regions [23, 24], and posterior variable selection summary [25]. Nonetheless, these approaches can only produce a single sparse model instead of several candidate models, and cannot generate credible sets similar to those obtained using discrete mixture priors.

In this paper, we introduce a novel fine-mapping method based on a continuous global-local shrinkage prior, called the heritability-induced Dirichlet decomposition (h2-D2) prior, which is a variant of R2-D2 prior [20]. R2-D2 prior possesses both unbounded density around the origin and very heavy tails, thus enabling it to model the extremely sparse structure of the fine-mapping coefficients. Our proposed h2-D2 prior inherits the same desirable properties as R2-D2 and is adapted specifically to GWAS data. Without loss of generality, we will refer to our method, which represents for the entire fine-mapping process, as h2-D2 throughout the manuscript.

Moreover, in order to address the limitations of continuous priors, inspired by the principles of frequentist hypothesis testing, we propose a statistic, termed “credible level”, which can be easily computed from posterior samples, to quantify how likely one or a set of SNPs have nonzero effects. We further define credible sets in the framework of continuous priors, offering a selection of candidate variants in the post-selection process.

Our simulation studies show that h2-D2 has better performance in identifying causal variants and accurately estimating effect sizes than the state-of-art fine-mapping methods such as SuSiE and FINEMAP. The CSs produced by h2-D2 exhibit superior power and achieve the target level of coverage when accurate LD matrices are provided. Finally, we apply h2-D2 to prostate cancer GWAS, identifying some novel causal signals that were not previously reported. The identified credible causal variants show significant enrichment in active gene regulatory regions and binding sites of specific transcription factors. In addition, we infer a total of 385 likely target genes associated with these credible causal variants. These genes are significantly over-represented in several pathways, providing valuable insights into the potential biological mechanisms underlying prostate cancer development and progression. We conclude with a discussion of future topics and further describe our software tool h2-D2 to implement the method for public use.

## Material and methods

### Overview of h2-D2

For a GWAS of quantitative trait with *N* individuals, consider a region containing *M* variants. The relationship between phenotypes and genotypes can be modeled by a multiple linear regression model:

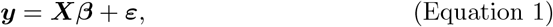

where ***y*** is a vector of standardized phenotype values for *N* individuals, ***X*** is an *N ×M* column-standardized genotype matrix for *N* individuals and *M* variants, ***β*** = (*β*_1_, …,*β*_*M*_)^*⊤*^ is an *M* -vector of effect sizes to be estimated, and ***ε*** is an *N* -vector of error terms. We assume that 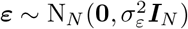, where ***I***_*N*_ is an *N × N* identity matrix and N_*k*_(***μ*, Λ**) denotes the *k*-variate normal distribution with mean ***μ*** and covariance matrix **Λ**.

We introduce a prior for ***β*** satisfying *E*(***β***) = **0** and Var(***β***) = **Σ**, where **Σ** is an *M × M* diagonal matrix with diagonal elements 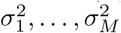. The narrow-sense heritability *h*^2^ of the quantitative trait explained by the *M* SNPs can be expressed as

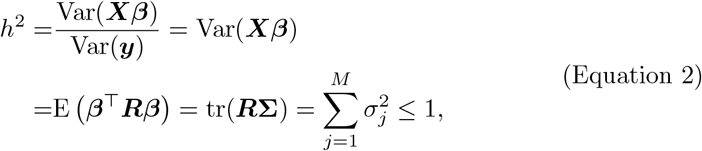

where ***R*** is the linkage disequilibrium (LD) matrix of the *M* variants. Then 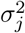 can be interpreted as the per-variant heritability of variant *j*.

To achieve an ideal prior that shrinks most elements of ***β*** toward 0 while retaining some large coefficients, we impose a Dirichlet prior on the variance terms:

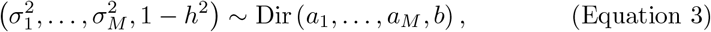

where *a*_1_, …, *a*_*M*_ ∈ (0, 1) and *b >* 0 are hyperparameters. Additionally, a double-exponential prior is assigned to each element of ***β***:

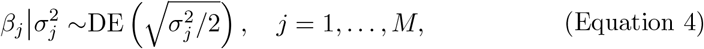

where DE(*δ*) denotes a double-exponential distribution with mean 0 and variance 2*δ*^2^.

Same as many other fine-mapping methods, h2-D2 requires GWAS summary data only [26]. Assume the single-SNP summary statistics 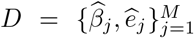 are provided, where 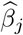 is the marginal effect size estimate o P *j*, and 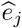 is its stan-dard error. Let 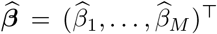,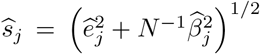 for *j* = 1, …, *M*, and 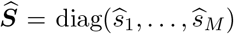. The LD matrix is estimated from some reference panel as 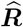. The RSS likelihood of 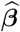 [26] is given by

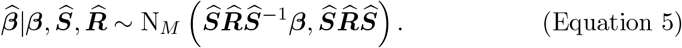

The h2-D2 prior can also be applied to binary traits by considering the observed-scale heritability (supplemental method 1). An MCMC algorithm that is compatible with both quantitative traits and binary traits is developed to obtain samples from the posterior distribution (supplemental method 2).

### Credible level and credible set

For the *j*-th SNP, consider the null hypothesis *H*_0*j*_ : *β*_*j*_ = 0. In the frequentist framework, *H*_0*j*_ can be rejected at the level of *α* if 0 is not contained in a confidence interval at the level of 1 −*α*. We migrate this approach to the Bayesian framework by replacing the confidence interval with the Bayesian credible interval. We propose the following statistic to evaluate how likely SNP *j* is causal:

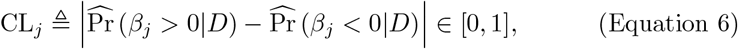

where the posterior probability 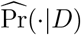 is estimated from the MCMC samples. We term this statistic as the “credible level” of SNP *j*, since it can be interpreted as the maximum probability such that the corresponding equal-tailed credible interval of *β*_*j*_ doesn’t cover 0.

Next, we extend this concept to multiple SNPs and define credible sets (CSs) accordingly. Consider a set of SNPs *C* = *{j*_1_, …, *j*_*k*_*}*. In the frequentist framework, claiming that *C* is a level 1 *− α* CS is equivalent to rejecting the null hypothesis 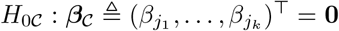 at the significance level of *α*, which can be declared if the null hypothesis ***v***^*⊤*^***β***_*𝒸*_ = 0 can be rejected at the significance level of *α* for at least one ***v*** *∈* ℝ^*k*^ . Therefore, the credible level of *𝒞* is defined as

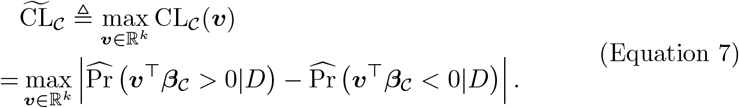

However, it is computationally infeasible to find the ***v*** that maximize the credible level when the number of variants exceeds two. Instead, we choose a single ***v*** to provide a lower bound of the credible level. The coefficients of ***v*** are selected to satisfy the condition that for positively correlated variants in 𝒞, the corresponding coeffi-cients have the same signs, while for negatively correlated variants, the corresponding coefficients have different signs. We choose ***v*** as an eigenvector of 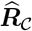 corresponding to its largest eigenvalue, where 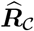 denotes the estimated LD matrix of SNPs in 𝒞. If CL_*C*_(***v***) ≥1−*α*, we have 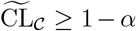, and we conclude that 𝒞 is a level 1−*α* credible set. A greedy algorithm is designed to search all CSs achieving a pre-specified level (supplemental method 3).

### Choice of hyper-parameters

In the Dirichlet prior (Equation 3), a smaller *a*_*j*_ leads to a higher concentration around 0 for *β*_*j*_, while a larger *b* indicates stronger global shrinkage. When incorporating external information, such as functional annotations, if the *j*-th SNP is more likely to be causal, a larger *a*_*j*_ can be set. By default, we suggest setting *a*_1_ = … = *a*_*M*_ = *a* ∈ [0.001, 0.01] for general fine-mapping tasks. A smaller *a* would make the MCMC chain converge slowly, while a larger *a* can be considered if there are evidences that the region may harbor a large number of causal variants (e.g. more than 10).

As for the choice of *b*, if an in-sample or highly accurate LD matrix 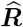 is available, we recommend estimating the local heritability using some well-known estimation procedures, such as the HESS estimator [27], which is defined as:

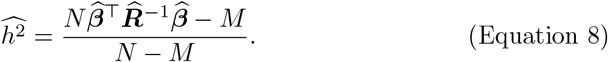

Then, *b* can be chosen as follows:

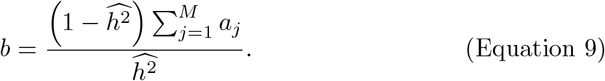

However, if the accuracy of 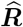 is poor, the HESS estimator may exhibit large bias. In this scenario, even if the true heritability is known, setting *b* according to Equation 9 can lead to large effect size estimates for some non-causal variants in h2-D2. This is consistent with a recent finding that significant miscalibration due to external LD matrices can produce suspicious results in meta-analysis fine-mapping studies [28]. To address this, we suggest performing quality-control to filter out outlier variants before fine-mapping, and setting 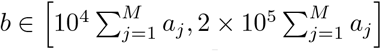 for GWAS fine-mapping tasks or setting 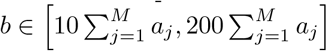 for eQTL fine-mapping tasks.

### UK Biobank data preprocessing

We selected British individuals from the UK Biobank database based on specific criteria. The selection process involved the following steps: (i) Only individuals with available genotype data were included. (ii) We specifically chose individuals who self-identified as “White British” to ensure homogeneity in the population. (iii) Genetic sex was confirmed to be consistent with self-reported sex. (iv) Outlier individuals were identified and excluded based on heterozygosity or missingness. (v) Individuals with close familial relationships were removed to avoid any potential bias in the analysis. After applying these filtering criteria, a total of 275, 768 individuals were retained for further analysis.

Subsequently, we focused on variants that met the following criteria: (i) Variants with at most one alternative allele were considered to ensure simplicity in the analysis. (ii) Variants with a minor allele frequency of at least 1% were selected to ensure a reasonable frequency of the variant in the population. (iii) Variants with an information score of at least 0.8 were included to ensure high-quality genotype data.. The rsID of selected variants were labeled based on dbSNP database (build 151).

### Partition LD blocks

We noticed that the LD blocks partitioned by LDetect [29] based on 1000 Genome reference panel are not optimal for UKBB reference panel. We developed a method to divide the whole genome into nearly independent LD blocks, so as to improve computational efficiency and achieve accurate fine-mapping results.

For a given LD matrix 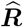 of *M* SNPs, we defined the optimal splitting as the solution to the following optimization problem:

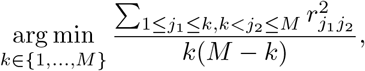

i.e., minimizing the average squared correlation *r*^2^ between two blocks. Our algorithm iteratively identifies optimal splitting points between consecutive LD blocks obtained from LDetect. If the loss in optimal splitting, defined as the difference in the objective function value before and after the split, is smaller than 0.001 and the size of the split block is not smaller than 50, the split point is accepted. This process is performed recursively for each split block until no further split points satisfying the conditions can be found.

As a result, we divided the entire autosomal region (excluding the major histocompatibility complex [MHC] regions) into a total of 3, 717 nearly independent LD blocks. We provide the script and the full list of LD blocks on our GitHub repository at https://github.com/xiangli428/PrCaFineMapping. This approach allows for improved efficiency and accuracy in fine-mapping analyses using the UK Biobank reference panel.

## Simulations

We conducted simulation studies using UK Biobank imputed genotype data from *N* = 275, 768 unrelated British individuals [30]. For the simulations, we selected 100 nearly independent LD blocks on chromosome 2 (Table S1), and included variants with MAF ≥0.01 and INFO score ≥0.8. We pruned SNPs such that the absolute correlation |*r*| between any two SNPs was less than 0.99. Each block contained a varying number of SNPs, ranging from 288 to 1, 122, and had a length between 0.25 and 2 Mb.

We designed four simulation scenarios with varying sample sizes, local heritabilities, and numbers causal variants. For the first three scenarios, we used the genotypes of all *N* = 275, 768 individuals and considered different combinations of local heritability and numbers of causal variants: (1) *h*^2^ = 0.1%, *n*_causal_ = 5; (2) *h*^2^ = 0.05%, *n*_causal_ = 5; and (3) *h*^2^ = 0.1%, *n*_causal_ = 10. In the last scenario, we simulated eQTL studies, where the sample sizes were small (*N* = 1, 000), but the effect sizes of causal SNPs were large (*h*^2^ = 10%), and *n*_causal_ = 5. Genotype values of each SNP were standardized. In each scenario, for each block, the causal variants were chosen randomly and the effect sizes of causal variants were sampled from a normal distribution with mean 0. The phenotype values were then computed according to the multiple regression model (Equation 1), where the error term ***ε*** were sampled from a multi-variate normal distribution with mean **0** and covariance matrix 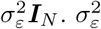 was chosen such that 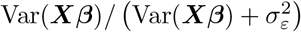 equaled *h*^*2*^ in each scenario. After standardizing the phenotype values and scaling the effect sizes of causal variants consistently, we computed summary statistics for each variant.

To assess the influence of LD matrix accuracy on the fine-mapping performance, we computed four LD matrics for each block. The first one was an in-sample LD matrix computed from all 275, 768 UKBB individuals 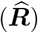 . The second one and the third one were down-sample LD matrices, computed from randomly sampled 3, 000 or 500 UKBB individuals, denoted by 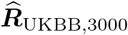 and 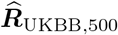, respectively. The fourth one was an out-of-sample LD matrix computed from 522 unrelated European ancestry individuals using the genotype data from the 1, 000 Genomes Project on GRCh38 [31, 32], denoted by 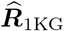. When using mismatched LD matrices, we applied SLALOM to all pairs of SNPs with |*r*| *≥* 0.8 and remove outlier non-causal variants with DENTIST-S statistics *≥* 40 [28].

### Compared methods

We performed a comprehensive comparison between h2-D2 and two state-of-art fine-mapping methods requiring only summary statistics, FINEMAP [10, 11] and SuSiE-RSS [12]. FINEMAP utilizes a general discrete distribution as prior for the number of causal SNPs,

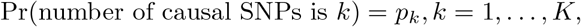

where *K ≪M* is the maximum number of causal variants, and uses a shotgun stochastic search algorithm to identify models with high posterior probabilities. SuSiE [6] is a novel variable selection method that decomposes the effect size as the sum of single-effect vectors and imposes a multinomial prior distribution on each single-effect vector. SuSiE adopts an iterative Bayesian stepwise selection algorithm to optimize a variational approximation to the posterior distribution, as well as a refinement procedure to address the convergence problem of the algorithm.

As for the choices of hyper-parameters, for h2-D2, we set *a*_1_ = … = *a*_*M*_ = *a* = 0.005. When using LD matrices 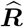 or 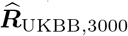, we set *b* according to Equation 8 and Equation 9. When using 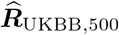 or 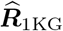, we set *b* = 2×10^5^*Ma* for scenario 1-3, and 100*Ma* for scenario 4, respectively. For SuSiE, we set options “refine=TRUE” and “estimate residual variance=TRUE”. For both SuSiE and FINEMAP, we set the number of single-effect vectors or the maximum number of causal variants equals the true number of causal variants in each scenario (5 or 10).

### Comparison of causal variant effect sizes and their posterior mean in simulation studies

In each simulation setting, we merged the results from 100 datasets together. Since causal variants with small effect sizes are difficult to be identified by fine-mapping methods, we used the following piecewise linear model to assess the relationship between the true effect sizes (*β*) of causal SNPs and their posterior means 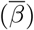:

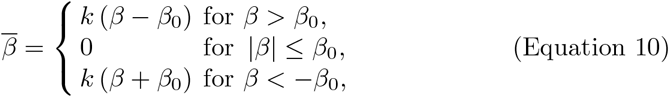

where *k* and *β*_0_ *>* 0 are the coefficients to be estimated. We used least square method to estimate these coefficients. The fitted curves are shown in Figure S4.

### Prostate cancer GWAS data preprocessing

We applied h2-D2 to identify candidate causal variants of prostate cancer (PrCa) using summary GWAS data from a large meta-analysis involving *N*_1_ = 79, 148 cases and *N*_0_ = 61, 106 controls of European ancestry [33]. We excluded the major histocompatibility complex MHC region (chr6 25-33 M) from our analysis. The remaining autosomal regions were partitioned into 3, 717 nonoverlapping regions with approximately independent LD (Material and methods). There are 126 risk variants out of 3, 717 regions (i.e., contain at least one SNP with *P <* 5×10^*−*8^). 275, 768 unrelated British individuals from UK Biobank database were used as reference panel.

We filtered out duplicated SNPs, SNPs that were not present in UKBB reference panel, SNPs with a imputation *r*^2^ *<* 0.3, with a standard error of marginal effect size on the allelic scale *<* 5×10^*−*3^ or *>* 10^*−*2^, with a MAF*<* 0.01 in UKBB reference panel, or with a logit(MAF) difference between UKBB reference panel and meta-analysis larger than 0.5. Since mismatched LD matrices were used, to avoid unreliable results, for each pair of SNPs with an absolute correlation |*r*| *≥*0.8, we checked if the pattern of LD and GWAS summary statistics is suspicious using DENTIST-S statistic [28]. If DENTIST-S statistic was greater than or equal to 30, the less significant SNP would be removed. After these quality control steps, 6, 446, 747 common SNPs were retained in our analysis. Before fine-mapping, the variants were pruned such that all pairwise correlation |*r*| *<* 0.95. A total of 1, 342, 667 tag SNPs were retained for fine-mapping. We used h2-D2 with specific hyper-parameters (*a*_1_ = … *aM* = 0.005 and *b* = 2×10^5^) to fine-map each region and identify 95% CSs. Each 95% CS includes a set of tag SNPs with a joint credible level *≥*0.95, as well as the pruned SNPs that are in high LD with them.

### Annotations of variants

The gene-based annotations of variants and their associated genes were extracted from the dbSNP database (build 151) with GRCh37.p13 as the reference assembly [34]. These annotations include: NSF (non-synonymous frameshift), NSM (non-synonymous missense), NSN (non-synonymous nonsense), SYN (synonymous), U3 (in 3’ UTR), U5 (in 5’ UTR), ASS (in acceptor splice site), DSS (in donor splice-site), INT (in intron), R3 (in 3’ gene region), and R5 (in 5’ gene region).

Prostate cancer-specific cis- and trans-eQTL data were obtained from PancanQTL database [35]. Cis-eQTLs from normal prostate tissues mapped in European-American subjects were obtained from GTEx V8 database [36].

DNaseI peaks, ChIP-seq peaks of histone marks and transcription factor binding sites in prostate-derived cell lines were obtained from Cistrome database [37]. Details of downloaded data are shown in Table S3. The peak coordinates were converted from hg38 to hg19 reference assembly using LiftOver. Variants located within these peaks were selected using BEDTools.

Enhancer-promoter loops identified from Hi-C data in RWPE1, C42B, and 22Rv1 cell lines were obtained from Supplementary Table 5A-C of ref. [38]. Annotated H3K27ac HiChIP loops in LNCaP cell line were obtained from Table S7 of ref. [39]. Variants located within the identified enhancers were selected using BEDTools.

### Pathway enrichment analysis

Potential target genes of credible causal variants (CCVs) were derived by merging (i) associated genes of CCVs annotated in dbSNP database (build 151); (ii) associated genes of eQTLs in CCVs in PancanQTL and GTEx V8 databases; (iii) genes whose promoters interact with enhancers covering CCVs in Hi-C or H3K27ac HiChIP data. Protein-coding genes were retained based on GENCODE v42 annotations mapped to GRCh37 assembly.

Enrichment analyses for pathways from GO Biological Process [40] and WikiPathways [41] were carried out using GeneCodis [42]. To remove redundant pathways, we computed Dice coefficients for all pairs of pathways. If the Dice coefficient between two pathways is larger than 0.3, only the more significant one was retained.

## Results

### Simulation results

We conducted simulation studies to evaluate the performance of h2-D2 and compared it with other fine-mapping methods. In brief, we chose 100 regions on chromosome 2 (Table S1) and simulated quantitative traits for each region. We considered four scenarios with varying sample sizes, local heritabilities, and numbers causal variants. To examine the influence of LD matrix accuracy on the fine-mapping performance, we computed four LD matrics from different reference panels with varying sample sizes for each block. Details are provided in Material and methods.

We compared h2-D2 with two state-of-art fine-mapping methods, FINEMAP [10, 11] and SuSiE-RSS [12]. On the SNP level, we evaluated the performance of variable selection using the area under the precision-recall curve (AUPRC), which was computed based on the credible level of each SNP for h2-D2 or the marginal posterior inclusion probability (PIP) of each SNP for SuSiE and FINEMAP. In addition, we assessed the accuracy of effect size estimation using the sum of squared error (SSE) of ***β*** based on its posterior mean. When using in-sample LD matrices, h2-D2 consistently outperformed SuSiE and FINEMAP in terms of both AUPRC and SSE across all scenarios (Figure 1A,B). As expected, all methods exhibited degraded performance as the accuracy of the LD matrices decreased. In most cases, h2-D2 still demonstrated superior performance. Additionally, h2-D2’s credible levels were better calibrated than PIPs of SuSiE and FINEMAP, particularly when inaccurate LD matrices were used (Figure S1). The performance of SuSiE was close to that of h2-D2. However, FINEMAP had significantly larger SSE and performed much worse in Scenario 3 where the true number of causal variants was 10.

**Figure 1.**
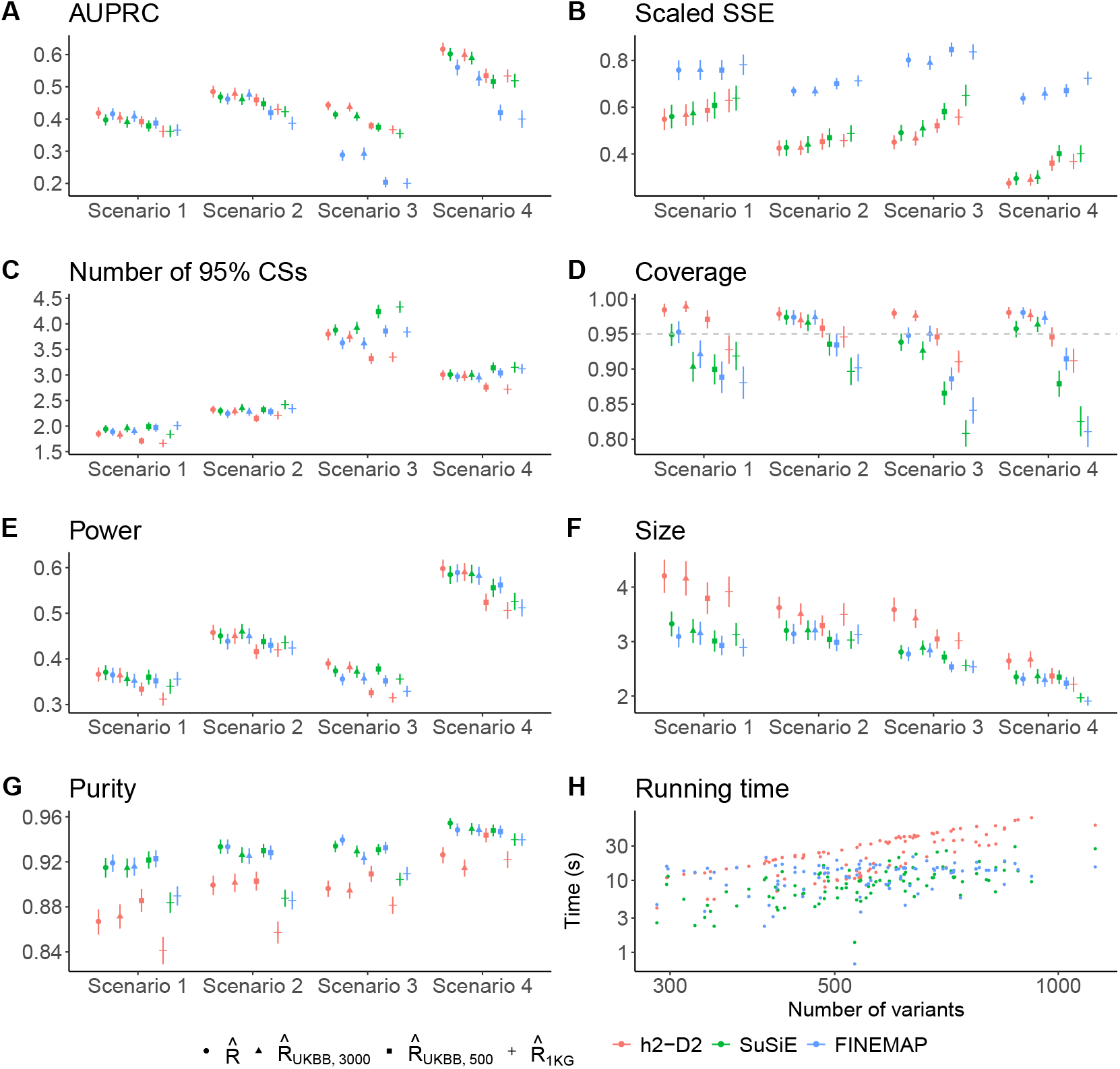
Performance comparison of h2-D2, SuSiE and FINEMAP on simulated data. In A-G, all values are the average ones across 100 datasets, with standard errors indicated by the error bars. (A) Area under the precision-recall curve (AUPRC) based on the credible level of each SNP for h2-D2 or the marginal posterior inclusion probability (PIP) of each SNP for SuSiE and FINEMAP. (B) Sum of squared error (SSE) of ***β*** based on its posterior mean, scaled by *h*^2^ in each scenario. (C) Number of detected 95% credible sets (CSs). (D) Coverage of 95% CS (the proportion of CSs that capture at least one causal variant). (E) Power of 95% CS (the proportion of causal variants captured by at least one CS). (F) Size of 95% CS (the number of variants in each CS). (G) Purity of 95% CS (the minimum absolute correlation among all pairs of SNPs in each CS). (H) Running time of the three methods against the number of variants in scenario 1. Each point represents a simulated dataset. For h2-D2, MCMC ran 10, 000 iterations. For SuSiE, *L* = 5. For FINEMAP, *K* = 5.

To gain further insights into the differences among the three methods, we compared the AUPRC for each simulated dataset between h2-D2 and the other two methods (Figure S2). While the AUPRC values were generally close for all three methods across most datasets, h2-D2 exhibited significantly better performance in certain datasets. By visualizing the fine-mapping results of these datasets, we noticed that in many cases if there was a non-causal SNP having moderate LD with one or more causal SNPs and having a stronger marginal association than causal SNPs, SuSiE and FINEMAP tended to select that non-causal SNP instead of the causal ones. Figure S3 provides two examples illustrating this issue. This phenomenon may be attributed to the stepwise selection nature of SuSiE and the shotgun stochastics search algorithm employed by FINEMAP. Once a marginally significant variant is included in the model, it is difficult for discrete-mixture prior-based methods to remove it, i.e., the algorithms are more prone to be trapped into suboptimal solutions. It appears that the refinement step of SuSiE cannot always alleviate this problem. On the other hand, continuous shrinkage prior-based methods allow for the continuous updating of coefficients, enabling smoother transitions among different local modes, and making the Markov Chain Monte Carlo (MCMC) algorithm to explore the space of causal configurations more extensively.

We also compared differences among the three methods in effect size estimation. We grouped the variants into causal and non-causal categories and analysed the prediction error for each group (Material and methods, Figures S4 and S5). Although SuSiE and h2-D2 produced similar estimation of causal variant effect sizes, h2-D2 had smaller prediction errors for the non-causal variant effect sizes, suggesting that h2-D2 had lower FDR than SuSiE. While FINEMAP demonstrated the lowest SSE for non-causal variant effect sizes, it grossly underestimated causal variant effect sizes, presumably from excessive shrinkage, resulting in larger SSE compared with SuSiE and h2-D2.

Next, we compared the level 95% CSs produced by the three methods. As shown in Figure 1C-G, when using 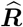 or 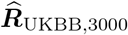, the numbers of 95% CSs generated by the three methods were comparable, and CSs from h2-D2 exhibited higher coverage and greater power in most cases. When using 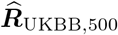 or 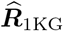, SuSiE and FINEMAP detected more CSs with higher power but lower coverage, while h2-D2 detected fewer CSs with lower power but higher coverage. These results suggested that the CSs from h2-D2 have a lower false discovery rate (FDR) even when low accuracy LD matrices are used. Although the CSs based on continuous priors may not guarantee the frequentist coverage, we found that the coverage was generally higher or close to the target level of 0.95, except when using 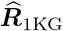. It is not surprising that 95% CSs from h2-D2 had larger sizes and lower purity, since SuSiE and FINEMAP focus on regions with high posterior probability density and select “best candidates” among a set of SNPs in high LD. In contrast, h2-D2 samples from the full posterior distribution, providing a more comprehensive representation of the uncertainty in the fine-mapping results.

Finally, we compared runtime of the three methods (Figure 1H). The computational complexity of h2-D2 is proportional to *M* ^2^ (where *M* is the number of variants) and the number of MCMC iterations (*n*_MCMC_), while the computational complexity of SuSiE is proportional to *M* ^2^ and the maximum number of single effects *L*. When *n*_MCMC_ = 10000 and *L* = 5, the runtime of h2-D2 were approximately three times as long as the runtime of SuSiE. The computational complexity of FINEMAP is primarily determined by the maximum number of causal variants and the number of iterations, so the runtime of FINEMAP didn’t significantly vary with the number of variants.

### Fine-mapping causal variants of prostate cancer

We applied h2-D2 to identify candidate causal variants of prostate cancer (PrCa) using summary GWAS data from a large meta-analysis of European ancestry [33] (Material and methods). Overall, we identified 164 CSs at 95% level (Table S2), containing 4, 706 credible causal variants (366 tags). Among these CSs, 93 overlaped with the 106 CSs in autosomal risk loci reported by ref. [43] and 86 overlaped with the CSs identified by ref. [39]. Out of the 3,717 regions analysed, 92 regions contained a single CS, while 23 regions contained multiple CSs. 6 CSs were detected within non-risk regions. The region with the highest number of CSs was chr8 127708268-128658961, where 15 CSs were detected. This finding is consistent with previous research that chr8q24 region harbors multiple loci associated with PrCa susceptibility [44]. The sizes of the CSs ranged from 1 to 282 variants, with a median size of 12 variants. There were 22 CSs containing only a single variant, including some well-established causal variants of PrCa, such as rs77559646, which disrupts *ANO7* mRNA splicing and protein expression [45], and rs61752561, which affects glycosylation and function of prostate-specific antigen [46] (Table 1).

**Table 1.**
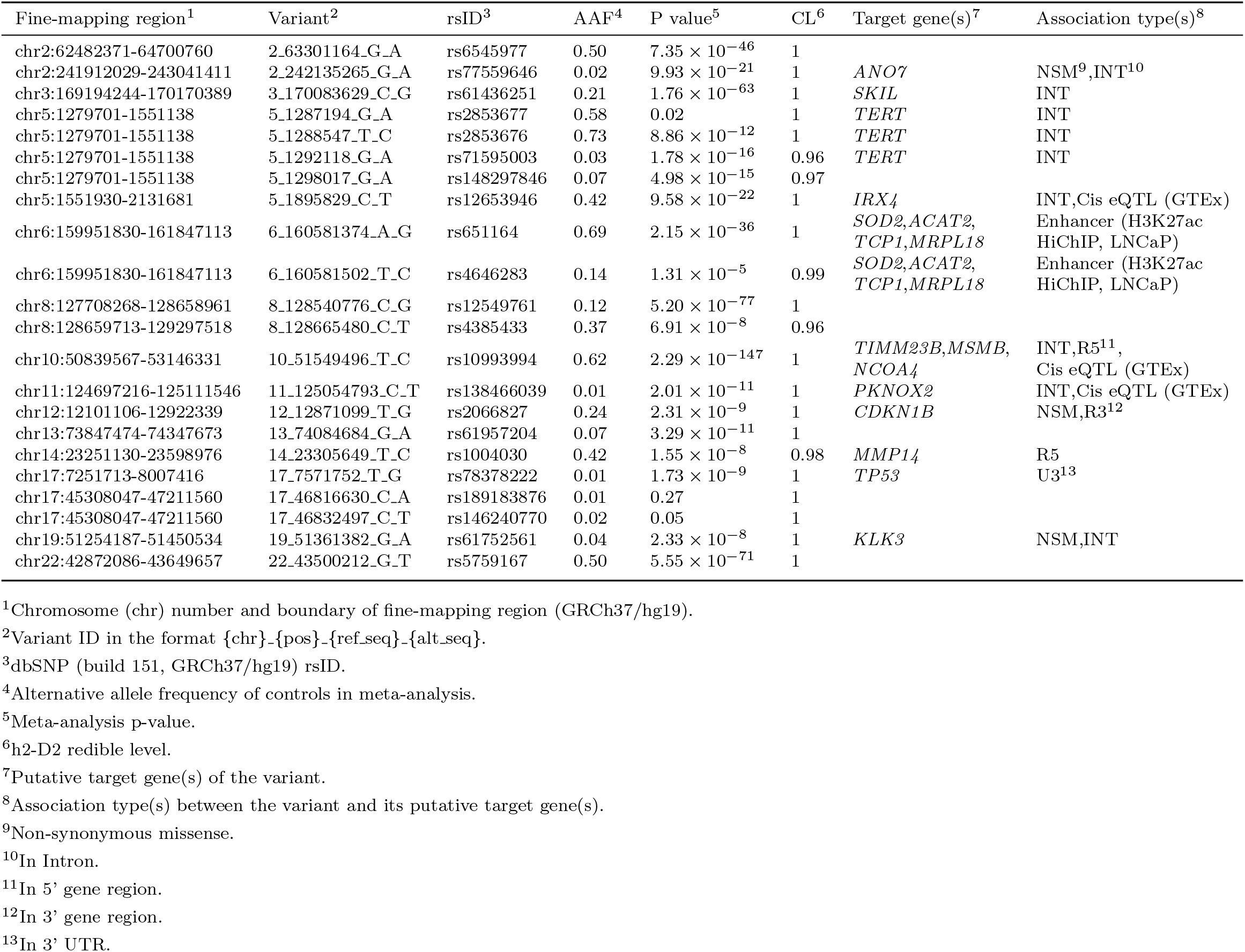
Single-SNP credible sets of prostate cancer causal variants.

In our analysis, we identified some novel independent association signals that have not been previously reported. One such example is chr11 68810837-69542062, where four 95% CSs were detected (Figure 2A, Figure S6A). CS:11-88-1 is represented by rs12275055 (*P* = 3.7×10^*−*98^), which is known to have pleiotropic associations with multiple cancer types [47]. This SNP acts as an eQTL in multiple tissues for *TPCN2*, which plays a role in autophagy progression and extracellular vesicle secretion in cancer cells [48]. The location of CS:11-88-2 overlaps with CS:11-88-1. Hi-C data from the normal prostate cell line RWPE1 indicated that several SNPs within CS:11-88-2 are located in an enhancer region that looping to the promoter of the cell cycle related gene *CCND1* [38]. Furthermore, an interaction between the *TPCN2* promoter and the *CCND1* promoter was detected by H3K27ac HiChIP in the LNCaP prostate cancer cell line [39]. These findings suggest a possible mechanism involving a three-way interaction between an enhancer harboring the causal SNPs in CS:11-88-1 and CS:11-88-2, the *TPCN2* promoter, and the *CCND1* promoter. We also identified two other CSs, CS:11-88-3 and CS:11-88-4, near the gene *CCND1*. Within CS:11-88-3, 4 out of 17 variants are located in the 5’ flanking region of *CCND1*. In CS:11-88-4, the most likely causal variant is the lead SNP rs3212870 (*P* = 1.5×10^*−*3^), which is located intronic in *CCND1*. The associations between CS:11-88-4 and PrCa have not been previously reported, because of the weak marginal associations, which can be explained by the moderate LD between CS:11-88-1, CS:11-88-2, and CS:11-88-4 (Figure S6A). Another interesting example is chr4 73256856-74885359 (Figure 2B, Figure S6B), where we identified a novel CS, CS:4-88-3, insignificantly associated with PrCa (minimum *P* = 6.5×10^*−*4^). The lead SNP in CS:4-88-3, rs72649118, is a non-synonymous missense SNP of *RASSF6*, a member of the RASSF family of tumor suppressors [49].

**Figure 2.**
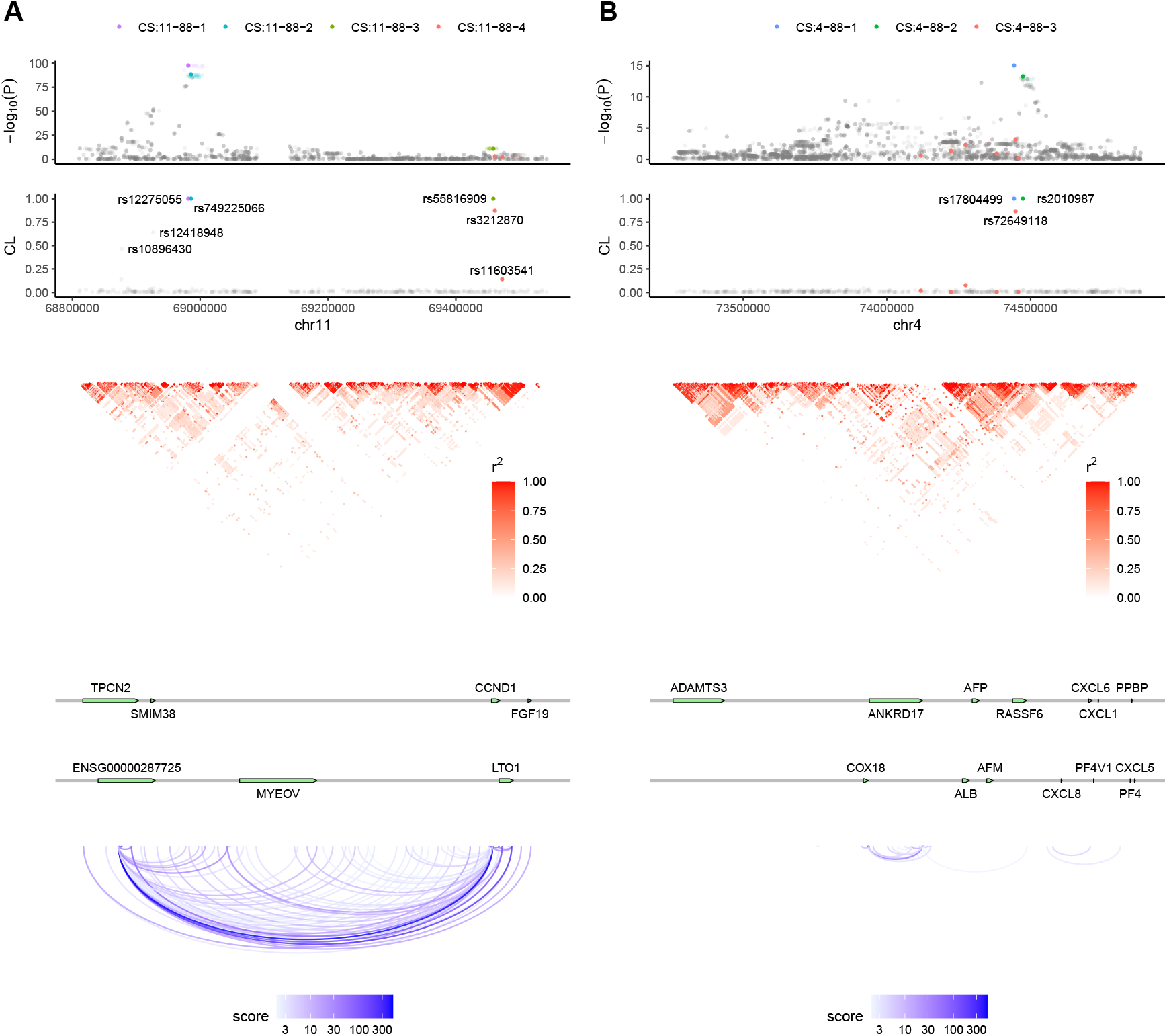
Fine-mapping results of two genomic regions in prostate cancer data analysis. (A) chr11 68810837-69542062; (B) chr4 73256856-74885359. The top panel depicts the marginal associations of variants (−log_10_(*P*)) from the GWAS meta-analysis data. The second panel illustrates the credible levels of tag SNPs computed by h2-D2. In the first two panels, each color represents a 95% credible set (CS). The CS is named in the format CS: {chromosome ID}-{region ID}-{index}. The third panel demonstrates the patterns of linkage disequilibrium of the genomic region. The fourth panel displays the positions of genes in the corresponding regions. The bottom panel shows the H3K27ac HiChIP loops detected in the LNCaP prostate cancer cell line [39].

### Functional enrichment of prostate cancer credible causal variants

We used the hypergeometric tests to investigate the enrichment of credible causal variants (CCVs) in specific genomic features, including prostate-specific DNaseI hyper-sensitivity sites, ChIP-seq peaks of transcription factors and histone marks (Material and methods, Table S3). We observed significant enrichment of CCVs in active regulatory regions (defined by H3K27ac and H3K4me1 marks), active gene promoters (defined by H3K4me3 and H3K9ac marks), actively transcribed gene bodies (defined by H3K36me3 and H3K79me2 marks), and DNaseI hypersensitivity sites (Figure 3A). CCVs were also significantly enriched in the binding sites of various transcription factors (Figure 3B, Table S3), including *AR* (androgen receptor), *NR3C1* (glucocorticoid receptor), *ASH2L*, and *FOXA1*.

**Figure 3.**
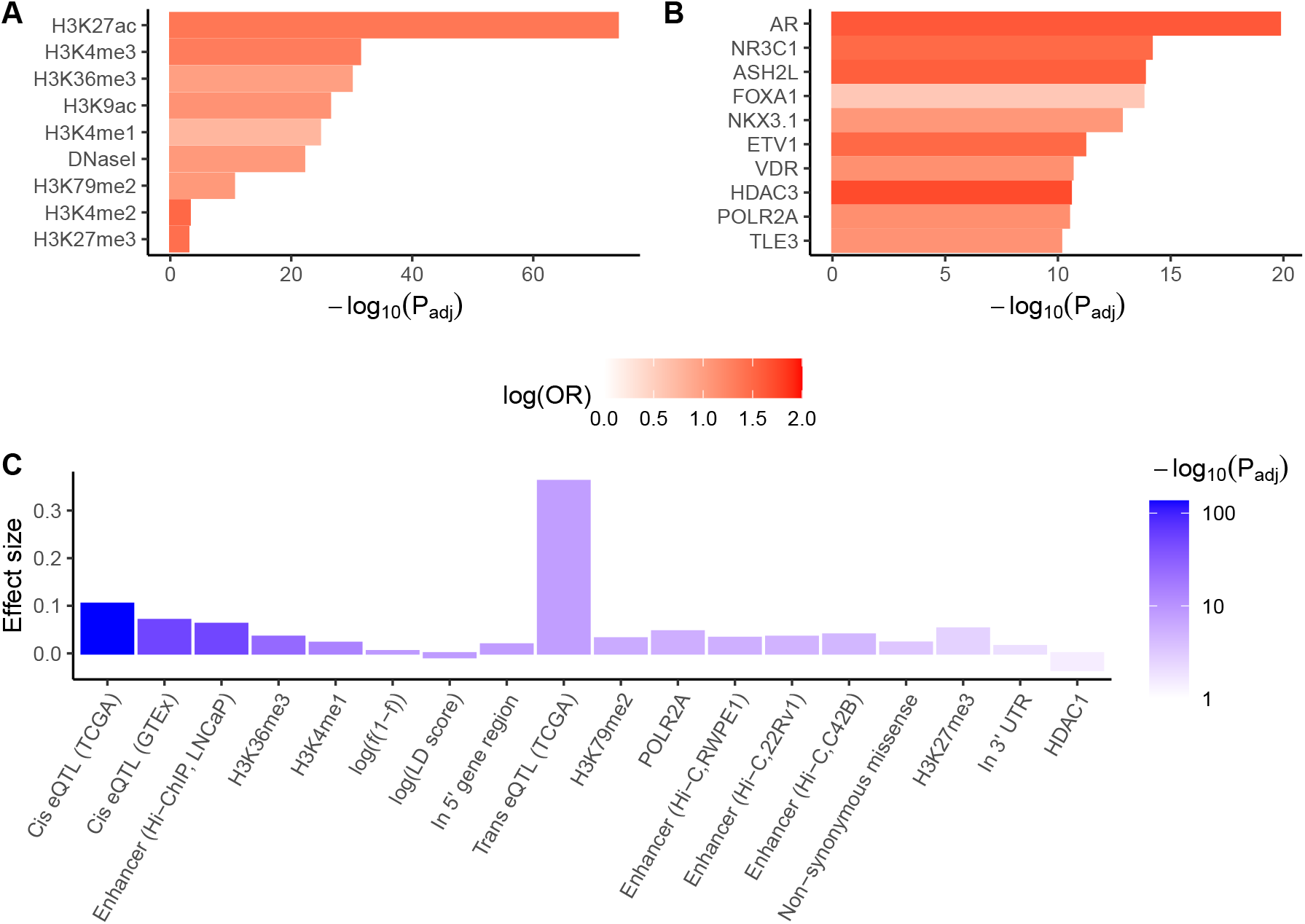
(A,B) Enrichment of credible causal variants in prostate-specific (A) histone marks and DNaseI hypersensitivity sites (B) top 10 transcription factor binding sites. Hypergeometric test *P* values are adjusted using the Benjamini-Hochberg (BH) method. (C) A linear regression model is fitted for the logarithm of per-SNP heritabilities of tag SNPs using the functional annotations of tag SNPs as predictors. Effect sizes and adjusted *P* values of significant functional annotations are shown. *P* values are adjusted using the BH method. Significance is defined as *P*_adj_ *<* 0.05.

To formally evaluate the relationship between the biological functions associated with SNPs and their contributions to the risk of PrCa, we fitted a linear model for the logarithm of per-SNP heritability (i.e., the posterior mean of squared effect size) of all 1, 342, 667 tag SNPs using the following functional annotations of SNPs as predictors: (i) 11 gene-based annotations extracted from the dbSNP database (build 151) [34]; (ii) cis- and trans-eQTLs within PrCa tissues from the TCGA database [35]; (iii) cis-eQTLs within normal prostate tissues from the GTEx v8 database [36]; (iv) DNaseI hypersensitivity sites, ChIP-seq peaks of 48 transcription factors and 9 histone modifications from normal prostate or prostate cancer cell lines, obtained from the Cistrome Data Browser [37]; (v) enhancer elements identified by Hi-C data and H3K27ac ChIP-seq peaks in normal prostate (RWPE1) and prostate cancer (C42B and 22Rv1) cell lines [38]; (vi) enhancer elements predicted by H3K27ac HiChIP in the prostate cancer cell line LNCaP [39]. In addition, log (*f* (1−*f*)) and log(LD score) were included as covariates, where *f* is the minor allele frequency of SNP. This analysis revealed that cis-eQTL (TCGA) (*P*_adj_ = 5.4×10^*−*137^), cis-eQTL (GTEx) (*P*_adj_ = 8.2×10^*−*52^), and enhancer (H3K27ac HiChIP, LNCaP) (*P*_adj_ = 1.8×10^*−*50^) were the most significant three annotations associated with per-SNP heritability (Figure 3C, Table S4). Trans-eQTL (TCGA) (*P*_adj_ = 1.5×10^*−*8^) exhibited the largest effect size (0.36). Notably, HDAC1 (histone deacetylase 1) binding site was the only significant functional annotation with a negative effect on per-SNP heritability. These findings suggested that genetic variants influencing gene expression levels and enhancer activity play a crucial role in the development and progression of PrCa.

### Putative target genes of prostate cancer credible causal variants

To identify potential target genes of CCVs, we integrated various sources of information, including gene-based annotations from the dbSNP database (build 151), eQTL data, and enhancer-promoter interaction data from Hi-C and HiChIP experiments (Material and methods). As a result, we identified 385 protein-coding genes as potential target genes of CCVs across all 95% CSs (Figure 4A, Table S2).

**Figure 4.**
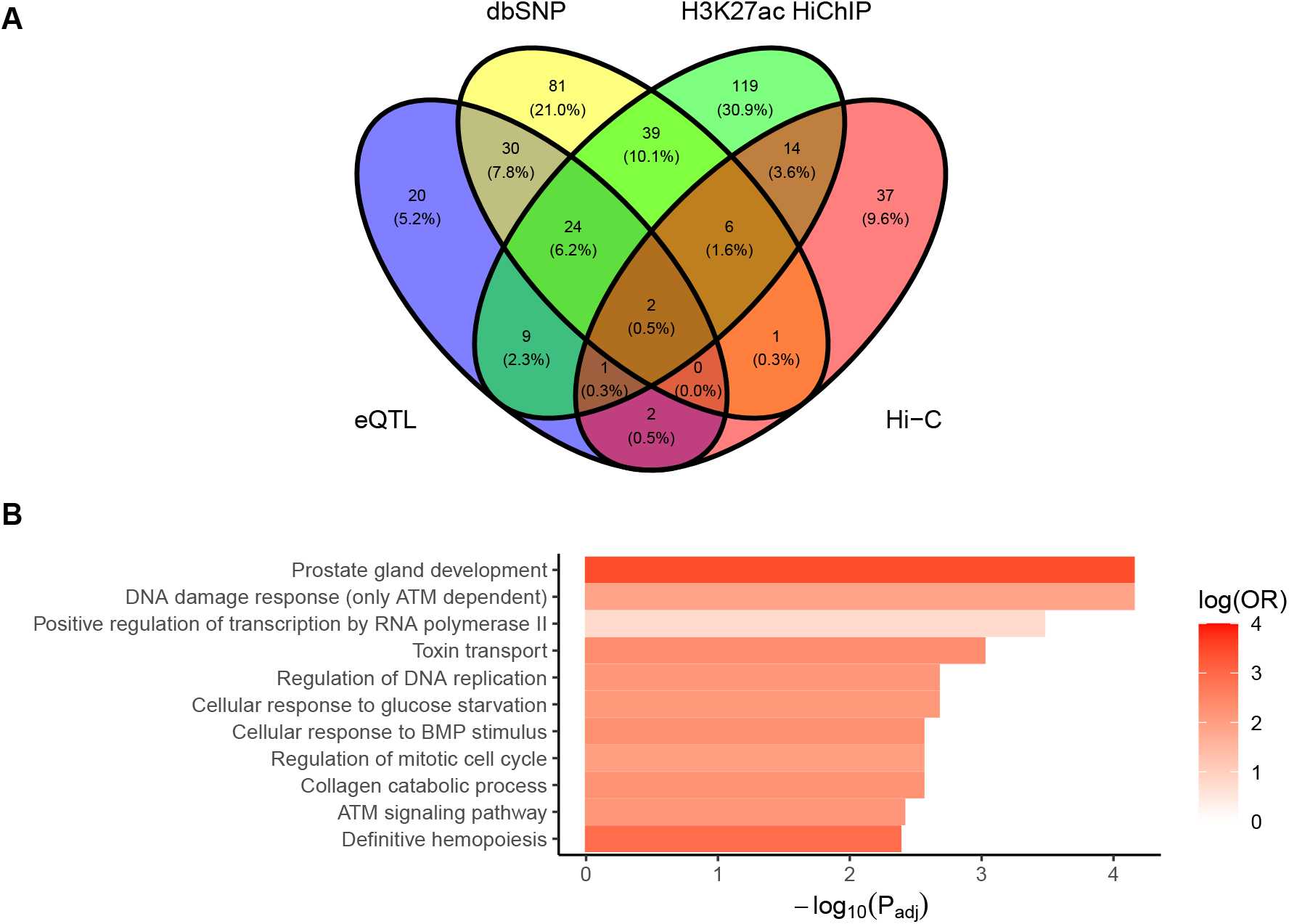
(A) Venn diagram showing the numbers of putative target genes inferred from different sources of information. (B) Enrichment of putative target genes in pathways from Gene Ontology Biological Processes and WikiPathways. Hypergeometric test *P* values are adjusted using the BH method. Pathways with *P*_adj_ *<* 0.005 are shown.

We further conducted pathway enrichment analysis to gain insights into the biological functions and processes associated with these putative target genes. Our analysis revealed significant over-representation of these genes in 52 non-redundant pathways at an FDR of 0.05 (Figure 4B, Table S5). Notable enriched pathways included prostate gland development, DNA damage response (only ATM dependent), positive regulation of transcription by RNA polymerase II, and regulation of mitotic cell cycle. The enrichment of putative target genes in cellular response to BMP (bone morphogenetic protein) stimulus, collagen catabolic process, and definitive hemopoiesis pathways may be attributed to the involvement of these processes in PrCa bone metastasis [50– 52]. Furthermore, putative target genes were also over-represented in toxin transport pathway. Although previous studies have reported associations between PrCa and several genes in toxin transport pathway, such as *SLC22A1–A3* [53, 54], the relationship between PrCa and this pathway is not well elucidated and needs further investigation.

## Discussion

In this article, we present h2-D2, a fine-mapping method that utilizes a continuous global-local shrinkage prior. As an extension of R2-D2, h2-D2 is designed for GWAS data where the phenotype values are standardized. Unlike existing fine-mapping methods that rely on discrete mixture priors, h2-D2 does not impose a constraint on the maximum number of causal variants and allows for the exploration of a wider range of causal configurations. In addition, h2-D2 does not reply on assumptions regarding the distribution of causal variant effect sizes, compatible with infinitesimal effect assumption for non-causal variants, which has been adopted by some recent works in fine-mapping [55, 56]. These features ensure the applicability and flexibility of h2-D2 in various scenarios.

We develop an efficient MCMC algorithm for h2-D2 to sample from the posterior distribution. We utilize several strategies to accelerate the mixing of MCMC chains, allowing for a more extensive exploration of the model space. Simulation studies show that h2-D2 is less likely to get trapped into local optima and performs better in variable selection than discrete-mixture-prior-based methods including SuSiE and FINEMAP. This may be due to the property of continuous priors that the coefficients are updated continuously and the transitions among local modes can be smoothly. Our results also highlight the importance of using accurate LD matrices derived from adequately large reference panels, which concurs with previous discoveries [12, 57].

Another important contribution of our work is that we propose an inference approach to define credible sets in the framework of continuous priors, which addresses the limitation of continuous priors that do not yield selection results directly. Simulation studies show that the CSs produced by h2-D2 can achieve the target level of coverage and are well-powered when using in-sample LD matrices, and exhibit an improved control of FDR when using mismatched LD matrices. These results suggest the robustness and effectiveness of our proposed approach. Theoretical properties of the credible level defined for multiple SNPs deserve further investigation. Additionally, we acknowledge that the greedy search algorithm used to identify credible sets may not always yield the optimal sets and may miss some sets (supplemental method 3). Further refinement and improvement of the algorithm are needed to enhance its performance.

In the real data application on prostate cancer GWAS, we identified several novel signals that have not been previously reported. Variants in 95 % CSs are significantly over-represented in prostate-specific epigenetic marks associated with activation of gene transcription. Through integrating gene-based annotation of SNPs, eQTL, Hi-C, and HiChIP data in prostate cell lines, we identified 385 potential target genes of variants in 95 % CSs. These genes are enriched in prostate development and cancer related pathways.

As a future direction, fine-mapping resolutions may be improved by integrating functional annotations into the h2-D2 prior. Stratified LD score regression-based methods like PolyFun [58] are well suited to be incorporated with h2-D2, since h2-D2 prior is imposed on the per-SNP heritability directly. Furthermore, h2-D2 can also be extended to multi-trait fine-mapping. Given the widespread existence of pleiotropy, fine-mapping multiple traits simultaneously has the potential to enhance the power of identifying shared causal variants among traits [59–61]. Jointly analyzing multiple traits may provide valuable insights into the genetic architecture underlying complex diseases and traits, and improve our understanding of the shared genetic basis between different phenotypes.

## Data Availability

All data produced in the present work are contained in the manuscript or are available online at https://github.com/xiangli428/PrCaFineMapping

https://www.ukbiobank.ac.uk/

https://www.internationalgenome.org/

http://practical.icr.ac.uk/blog/?page_id=8164

https://ftp.ncbi.nih.gov/snp/organisms/human_9606_b151_GRCh37p13/

http://gong_lab.hzau.edu.cn/PancanQTL/

https://gtexportal.org/home/

http://cistrome.org/

https://static-content.springer.com/esm/art%3A10.1038%2Fs41467-019-12079-8/MediaObjects/41467_2019_12079_MOESM7_ESM.xlsx

https://ars.els-cdn.com/content/image/1-s2.0-S0002929721004195-mmc3.csv

https://github.com/xiangli428/PrCaFineMapping

## Data and code availability

Prostate cancer summary data are available from the PRACTICAL Consortium (http://practical.icr.ac.uk/blog/?page_id=8164). Enhancer-promoter loops identified from Hi-C data in RWPE1, C42B, and 22Rv1 cell lines are available at https://static-content.springer.com/esm/art%3A10.1038%2Fs41467-019-12079-8/MediaObjects/41467_2019_12079_MOESM7_ESM.xlsx. Annotated H3K27ac HiChIP loops in LNCaP cell line are available at https://ars.els-cdn.com/content/image/1-s2. 0-S0002929721004195-mmc3.csv. The software h2D2 is available at https://github.com/xiangli428/h2D2. Scripts and data related to PrCa fine-mapping analysis are available at https://github.com/xiangli428/PrCaFineMapping.

## Acknowledgments

This work was supported, in part, by the Hong Kong Research Grants Council (RGC) Early Career Scheme 2021/22 (project number 27305221).

## Web resources

UK Biobank, https://www.ukbiobank.ac.uk/

1000 Genomes on GRCh38, https://www.internationalgenome.org/

Prostate cancer summary data, http://practical.icr.ac.uk/blog/?page_id=8164

dbSNP (build 151) with GRCh37.p13 as reference assembly, https://ftp.ncbi.nih.gov/snp/organisms/human_9606_b151_GRCh37p13/.

PancanQTL, http://gong_lab.hzau.edu.cn/PancanQTL/

GTEx_V8, https://gtexportal.org/home/

Cistrome, http://cistrome.org/

plink, https://zzz.bwh.harvard.edu/plink/

BEDTools, https://bedtools.readthedocs.io/en/latest/

LDetect, https://bitbucket.org/nygcresearch/ldetect/src/master/

bigsnpr, https://privefl.github.io/bigsnpr/

GeneCodis, https://genecodis.genyo.es/

FINEMAP, http://christianbenner.com/SuSiE, https://github.com/stephenslab/susieR

During the preparation of this work the authors used ChatGPT in order to improve the expression. After using this tool/service, the authors reviewed and edited the content as needed and take full responsibility for the content of the publication.

