## Supplementary Note and Figures for "A novel Bayesian fine-mapping model using a continuous global-local shrinkage prior with applications in prostate cancer analysis" (from another version of this preprint: version 1, 2023-08-08)

#### 1 Binary traits

##### 1.1 First order approximation for logistic model

For binary traits, we consider the following logistic regression model:

$$p_i = \Pr\left(y_i = 1 \middle| x_i, \beta_0, \boldsymbol{\beta}\right) = \frac{\exp\left(\beta_0 + \boldsymbol{x}_i^\top \boldsymbol{\beta}\right)}{1 + \exp\left(\beta_0 + \boldsymbol{x}_i^\top \boldsymbol{\beta}\right)}, i = 1, \ldots, N, \tag{1}$$

where $y_i \in \{0, 1\}$ is the phenotype value of individual $i$, $y_i = 1$ for cases and $y_i = 0$ for controls. $x_i$ is the genotype vector of individual $i$. $\beta_0$ is the intercept, and $\boldsymbol{\beta}$ is the effect size vector of $M$ SNPs.

Let $\boldsymbol{y} = (y_1, \ldots, y_N)^\top$ be the phenotype vector, and $\boldsymbol{X} = (\boldsymbol{x}_1, \ldots, \boldsymbol{x}_N)^\top$ be the genotype matrix. Each column of $\boldsymbol{X}$ is standardized with mean 0 and variance 1. Denote the number of cases as $N_1$, and the number of controls as $N_0$. $N_1 + N_0 = N$. Let $\widetilde{\boldsymbol{\beta}} = \left(\beta_0, \boldsymbol{\beta}^\top\right)^\top$, $\widetilde{\boldsymbol{x}}_i = \left(1, \boldsymbol{x}_i^\top\right)^\top$, $\widetilde{\boldsymbol{X}} = [\mathbf{1}_N, \boldsymbol{X}]$, and $\boldsymbol{p} = (p_1, \ldots, p_N)^\top$. The log-likelihood function can be expressed as

$$\begin{aligned} L\left(\widetilde{\boldsymbol{\beta}}\right) &= \sum_{i=1}^{N} \left[y_i \log p_i + (1 - y_i) \log(1 - p_i)\right] \\ &= \boldsymbol{y}^\top \widetilde{\boldsymbol{X}} \widetilde{\boldsymbol{\beta}} - \sum_{i=1}^{N} \log\left(1 + \exp\left(\widetilde{\boldsymbol{x}}_i^\top \widetilde{\boldsymbol{\beta}}\right)\right). \end{aligned} \tag{2}$$

The score function is

$$\frac{\partial L}{\partial \widetilde{\boldsymbol{\beta}}} = \widetilde{\boldsymbol{X}}^\top (\boldsymbol{y} - \boldsymbol{p}). \tag{3}$$

We adopt the first order approximation derived by[1] to deal with the score function. Under the small-effect assumption, $\left|\boldsymbol{x}_i^\top \boldsymbol{\beta}\right|$ is small. By expanding $p_i$ as a Taylor series around $\left|\boldsymbol{x}_i^\top \boldsymbol{\beta}\right| = 0$, we have

$$\begin{aligned} p_i &= \frac{\mathrm{e}^{\beta_0}}{1+\mathrm{e}^{\beta_0}} + \frac{\mathrm{e}^{\beta_0}}{\left(1+\mathrm{e}^{\beta_0}\right)^2} \boldsymbol{x}_i^\top \boldsymbol{\beta} + o\left(\left(\boldsymbol{x}_i^\top \boldsymbol{\beta}\right)^2\right) \\ &\approx \gamma_0 + \boldsymbol{x}_i^\top \boldsymbol{\gamma} = \widetilde{\boldsymbol{x}}_i^\top \widetilde{\boldsymbol{\gamma}}, \end{aligned} \tag{4}$$

where $\gamma_0 = \mathrm{e}^{\beta_0} / \left(1+\mathrm{e}^{\beta_0}\right)$, $\boldsymbol{\gamma} = \gamma_0 \left(1-\gamma_0\right) \boldsymbol{\beta}$, and $\widetilde{\boldsymbol{\gamma}} = \left(\gamma_0, \boldsymbol{\gamma}^\top\right)^\top$. The score function can be approximated as

$$\frac{\partial L}{\partial \widetilde{\boldsymbol{\beta}}} \approx \widetilde{\boldsymbol{X}}^\top \boldsymbol{y} - \widetilde{\boldsymbol{X}}^\top \widetilde{\boldsymbol{X}} \widetilde{\boldsymbol{\gamma}}, \tag{5}$$

which corresponds to the linear model

$$\boldsymbol{y} = \gamma_0 + \boldsymbol{X}\boldsymbol{\gamma}. \tag{6}$$

The least squares estimate of $\widetilde{\boldsymbol{\gamma}}$ is

$$\check{\gamma}_0 = \frac{N_1}{N}, \check{\boldsymbol{\gamma}} = \left(\boldsymbol{X}^\top \boldsymbol{X}\right)^{-1} \boldsymbol{X}^\top \boldsymbol{y}. \tag{7}$$

In the following derivation, we fix $\gamma_0$ as $N_1/N$. The least squares estimate of $\widetilde{\boldsymbol{\beta}}$ becomes

$$\check{\beta}_0 = \log\left(\frac{N_1}{N_0}\right), \check{\boldsymbol{\beta}} = \frac{N^2}{N_1 N_0} \left(\boldsymbol{X}^\top \boldsymbol{X}\right)^{-1} \boldsymbol{X}^\top \boldsymbol{y} = c^{-2} \left(\boldsymbol{X}^\top \boldsymbol{X}\right)^{-1} \boldsymbol{X}^\top \boldsymbol{y}, \tag{8}$$

where $c^2 = N_1 N_0 / N^2 = \mathrm{Var}(\boldsymbol{y})$.

##### 1.2 h2-D2 prior for binary traits

According to the linear model (6), the observed-scale heritability can be expressed as

$$\begin{aligned} h_o^2 &= \frac{\mathrm{Var}(\boldsymbol{X}\boldsymbol{\gamma})}{\mathrm{Var}(\boldsymbol{y})} = \frac{c^4 \,\mathrm{Var}(\boldsymbol{X}\boldsymbol{\beta})}{c^2} \\ &= c^2 \,\mathrm{Var}(\boldsymbol{X}\boldsymbol{\beta}) = c^2 \mathrm{E}\left(\boldsymbol{\beta}^\top \boldsymbol{R} \boldsymbol{\beta}\right) = c^2 \,\mathrm{tr}(\boldsymbol{R}\boldsymbol{\Sigma}) = c^2 \sum_{j=1}^{M} \sigma_j^2. \end{aligned} \tag{9}$$

Therefore we assign the following Dirichlet decomposition prior to the observed-scale heritability.

$$\left(c^2\sigma_1^2, \ldots, c^2\sigma_M^2, 1-h_o^2\right) \sim \ \mathrm{Dir}\left(a_1, \ldots, a_M, b\right). \tag{10}$$

##### 1.3 Likelihood of GWAS summary data for binary traits

Under the small-effect assumption, according to Equation (8), the marginal effect size of SNP $j$ estimated by logistic regression model can be approximated as

$$\widehat{\beta}_j \approx \frac{N}{N_1 N_0} \boldsymbol{X}_j^\top \boldsymbol{y}, \tag{11}$$

with approximated variance

$$\operatorname{Var}\left(\widehat{\beta}_j\right) \approx \frac{N}{N_1 N_0}. \tag{12}$$

Other test statistics, such as Armitage's trend test statistic and score test statistic, have similar form as this estimate[2].

Let $\widehat{\boldsymbol{\beta}} = (\widehat{\beta}_1, \ldots, \widehat{\beta}_M)^\top$. To derive the joint distribution of $\widehat{\boldsymbol{\beta}}$ condition on $\boldsymbol{\beta}$, we expand the log-likelihood function (2) as

$$\begin{aligned}
L\left(\widetilde{\boldsymbol{\beta}}\right) &= \boldsymbol{y}^\top \widetilde{\boldsymbol{X}} \widetilde{\boldsymbol{\beta}} - \sum_{i=1}^N \log\left(1 + \exp\left(\widetilde{\boldsymbol{x}}_i^\top \widetilde{\boldsymbol{\beta}}\right)\right) \\
&\approx N_1 \beta_0 + \boldsymbol{\beta}^\top \boldsymbol{X}^\top \boldsymbol{y} - \sum_{i=1}^N \left[\log\left(1 + \mathrm{e}^{\beta_0}\right) + \frac{\mathrm{e}^{\beta_0}}{1 + \mathrm{e}^{\beta_0}} \boldsymbol{x}_i^\top \boldsymbol{\beta} + \frac{\mathrm{e}^{\beta_0}}{2\left(1 + \mathrm{e}^{\beta_0}\right)^2} \left(\boldsymbol{x}_i^\top \boldsymbol{\beta}\right)^2\right] \\
&= C + \boldsymbol{\beta}^\top \boldsymbol{X}^\top \boldsymbol{y} - \frac{\mathrm{e}^{\beta_0}}{2\left(1 + \mathrm{e}^{\beta_0}\right)^2} \boldsymbol{\beta}^\top \boldsymbol{X}^\top \boldsymbol{X} \boldsymbol{\beta} \\
&\approx C + \frac{N_1 N_0}{N} \boldsymbol{\beta}^\top \widehat{\boldsymbol{\beta}} - \frac{1}{2} \frac{N_1 N_0}{N} \boldsymbol{\beta}^\top \widehat{\boldsymbol{R}} \boldsymbol{\beta},
\end{aligned} \tag{13}$$

given that $\sum_{i=1}^N \boldsymbol{x}_i = \mathbf{0}$ and $\sum_{i=1}^N \boldsymbol{x}_i \boldsymbol{x}_i^\top = \boldsymbol{X}^\top \boldsymbol{X} = N \widehat{\boldsymbol{R}}$, since $\boldsymbol{X}$ is standardized. Therefore, the distribution of $\widehat{\boldsymbol{\beta}}$ condition on $\boldsymbol{\beta}$ can be approximated as

$$\widehat{\boldsymbol{\beta}} | \boldsymbol{\beta}, \widehat{\boldsymbol{R}} \sim \mathrm{N}\left(\widehat{\boldsymbol{R}} \boldsymbol{\beta}, \frac{N}{N_1 N_0} \widehat{\boldsymbol{R}}\right). \tag{14}$$

If we set $\boldsymbol{S} = \sqrt{\frac{N}{N_1 N_0}} I_M$, we will get a same form as RSS likelihood for quantitative traits[3]. In application, for binary traits, we set $\widehat{s}_j$ as the standard error of $\widehat{\beta}_j$ for $j = 1, \ldots, M$, and plug in the estimate $\widehat{\boldsymbol{S}} = \operatorname{diag}(\widehat{s}_1, \ldots, \widehat{s}_M)$ for $\boldsymbol{S}$.

#### 2 MCMC sampling

For $j = 1, \ldots, M$, we introduce a latent variable $\psi_j \sim \operatorname{Exp}(1)$, such that the Laplace prior distribution $\beta_j | \sigma_j^2 \sim \operatorname{DE}\left(0, \sqrt{\sigma_j^2 / 2}\right)$ can be expressed as

$$\begin{aligned}\beta_j \big| \psi_j, \sigma_j^2 \sim & \mathrm{N}\left(0, \psi_j \sigma_j^2\right), \quad j = 1, \ldots, M, \\ \psi_j \sim & \mathrm{Exp}(1), \quad j = 1, \ldots, M.\end{aligned}$$

We adopt the following reparametrization for $\boldsymbol{\sigma}^2$. $\tau_j = \log\left(c^2\sigma_j^2\right)$, $j = 1, \ldots, M$, where $c^2 = 1$ for quantitative traits and $c^2 = N_1 N_0 / N^2$ for binary traits. The prior probability distribution for $\boldsymbol{\tau} = (\tau_1, \ldots, \tau_M)$ is

$$p(\tau_1, \ldots, \tau_M) \propto \prod_{j=1}^{M} \mathrm{e}^{a_j \tau_j} \times \left(1 - \sum_{j=1}^{M} \mathrm{e}^{\tau_j}\right)^{b-1} \times \mathbf{1}\left\{\sum_{j=1}^{M} \mathrm{e}^{\tau_j} \leq 1\right\}. \tag{15}$$

The complete data likelihood is

$$\begin{aligned}& p\left(\widehat{\boldsymbol{\beta}}, \boldsymbol{\beta}, \boldsymbol{\tau}, \boldsymbol{\psi} \big| \widehat{\boldsymbol{S}}, \widehat{\boldsymbol{R}}\right) \\ \propto & \mathcal{N}_M\left(\widehat{\boldsymbol{\beta}}; \widehat{\boldsymbol{S}}\widehat{\boldsymbol{R}}\widehat{\boldsymbol{S}}^{-1}\boldsymbol{\beta}, \widehat{\boldsymbol{S}}\widehat{\boldsymbol{R}}\widehat{\boldsymbol{S}}\right) \\ \times & \prod_{j=1}^{M} \mathcal{N}_1\left(\beta_j; 0, \psi_j \mathrm{e}^{\tau_j}/c^2\right) \times \prod_{j=1}^{M} \exp\left(-\psi_j\right) \\ \times & \prod_{j=1}^{M} \mathrm{e}^{a_j \tau_j} \times \left(1 - \sum_{j=1}^{M} \mathrm{e}^{\tau_j}\right)^{b-1} \times \mathbf{1}\left\{\sum_{j=1}^{M} \mathrm{e}^{\tau_j} \leq 1\right\},\end{aligned}$$

where $\mathcal{N}_k$ is the probability density function of $k$-variate normal distribution. Notice that

$$\begin{aligned}& \mathcal{N}_M\left(\widehat{\boldsymbol{\beta}}; \widehat{\boldsymbol{S}}\widehat{\boldsymbol{R}}\widehat{\boldsymbol{S}}^{-1}\boldsymbol{\beta}, \widehat{\boldsymbol{S}}\widehat{\boldsymbol{R}}\widehat{\boldsymbol{S}}\right) \\ \propto & \exp\left\{-\frac{1}{2}\left(\widehat{\boldsymbol{\beta}} - \widehat{\boldsymbol{S}}\widehat{\boldsymbol{R}}\widehat{\boldsymbol{S}}^{-1}\boldsymbol{\beta}\right)^{\top} \widehat{\boldsymbol{S}}^{-1}\widehat{\boldsymbol{R}}^{-1}\widehat{\boldsymbol{S}}^{-1}\left(\widehat{\boldsymbol{\beta}} - \widehat{\boldsymbol{S}}\widehat{\boldsymbol{R}}\widehat{\boldsymbol{S}}^{-1}\boldsymbol{\beta}\right)\right\} \\ \propto & \exp\left\{-\frac{1}{2}\boldsymbol{\beta}^{\top}\widehat{\boldsymbol{S}}^{-1}\widehat{\boldsymbol{R}}\widehat{\boldsymbol{S}}^{-1}\boldsymbol{\beta} + \widehat{\boldsymbol{\beta}}^{\top}\widehat{\boldsymbol{S}}^{-2}\boldsymbol{\beta}\right\},\end{aligned}$$

we propose the following MCMC sampling algorithm without computing the inverse of LD matrix.

(a) Set initial values for $\boldsymbol{\tau}$, $\boldsymbol{\beta}$, and $\boldsymbol{\psi}$.

(b) For $j = 1, \ldots, M$, update $\tau_j$ using MH algorithm according to the unnormalized density

$$\begin{aligned}& p\left(\tau_j \big| \boldsymbol{\beta}_{-j}, \psi_j, \boldsymbol{\tau}_{-j}, \widehat{\beta}_j, \widehat{\boldsymbol{S}}, \widehat{\boldsymbol{R}}\right) \\ \propto & \widetilde{\sigma}_j \times \exp\left\{\frac{1}{2}u_j^2\widetilde{\sigma}_j^2\right\} \times \mathrm{e}^{\tau_j\left(a_j - \frac{1}{2}\right)} \\ \times & \left(1 - \sum_{k \neq j} \mathrm{e}^{\tau_k} - \mathrm{e}^{\tau_j}\right)^{b-1} \times \mathbf{1}\left\{\mathrm{e}^{\tau_j} \leq 1 - \sum_{k \neq j} \mathrm{e}^{\tau_k}\right\},\end{aligned} \tag{16}$$

where

$$\widetilde{\sigma}_j^2 = \left[\widehat{s}_j^{-2} + \frac{c^2}{\psi_j \mathrm{e}^{\tau_j}}\right]^{-1}, u_j = \widehat{s}_j^{-2}\widehat{\beta}_j - \sum_{k\neq j} \widehat{\boldsymbol{W}}_{kj}\beta_k,$$

and $\widehat{\boldsymbol{W}} = \widehat{\boldsymbol{S}}^{-1}\widehat{\boldsymbol{R}}\widehat{\boldsymbol{S}}^{-1}$.

We apply the following jumping rule to purpose a candidate for $\tau_j$:

$$\tau_j^* | \tau_j \sim \log\left(1 - \sum_{k\neq j} \mathrm{e}^{\tau_k}\right) + \left[\tau_j - \log\left(1 - \sum_{k\neq j} \mathrm{e}^{\tau_k}\right)\right] \times \mathrm{N}(0, \delta^2),$$

and accept $\tau_j^*$ with probability

$$\begin{aligned} &\frac{p\left(\tau_j^* | \boldsymbol{\beta}_{-j}, \psi_j, \boldsymbol{\tau}_{-j}, \widehat{\beta}_j, \widehat{\boldsymbol{S}}, \widehat{\boldsymbol{R}}\right)}{p\left(\tau_j | \boldsymbol{\beta}_{-j}, \psi_j, \boldsymbol{\tau}_{-j}, \widehat{\beta}_j, \widehat{\boldsymbol{S}}, \widehat{\boldsymbol{R}}\right)} \times \frac{q\left(\tau_j | \tau_j^*\right)}{q\left(\tau_j^* | \tau_j\right)} \\ &= \left(\frac{c^2 + \widehat{s}_j^{-2}\psi_j \mathrm{e}^{\tau_j}}{c^2 + \widehat{s}_j^{-2}\psi_j \mathrm{e}^{\tau_j^*}}\right)^{1/2} \times \exp\left\{\frac{u_j^2}{2}\left(\widetilde{\sigma}_j^{*2} - \widetilde{\sigma}_j^2\right)\right\} \times \mathrm{e}^{a_j\left(\tau_j^* - \tau_j\right)} \\ &\times \left(\frac{1 - \sum_{k\neq j} \mathrm{e}^{\tau_k} - \mathrm{e}^{\tau_j^*}}{1 - \sum_{k\neq j} \mathrm{e}^{\tau_k} - \mathrm{e}^{\tau_j}}\right)^{b-1} \times \frac{\tau_j^* - \log\left(1 - \sum_{k\neq j} \mathrm{e}^{\tau_k}\right)}{\tau_j - \log\left(1 - \sum_{k\neq j} \mathrm{e}^{\tau_k}\right)}. \end{aligned} \tag{17}$$

In application, we set the the default stepsize $\delta = 2$ such that the acceptance rate is around 0.44.

(c) For $j = 1, \ldots, M$, update $\beta_j \sim \mathrm{N}(\widetilde{\sigma}_j^2 u_j, \widetilde{\sigma}_j^2)$.

(d) For $j = 1, \ldots, M$, update

$$\psi_j^{-1} | \beta_j, \tau_j \sim \mathrm{InvGaussian}\left(\mu = \frac{\mathrm{e}^{\tau_j/2}}{c\,|\beta_j|}, \lambda = 1\right).$$

(e) To improve mixing of MCMC chain, after every 10 steps, we will choose a pair of SNPs with high LD, $j_1$ and $j_2$, and propose switching the values of $\tau_{j_1}$ and $\tau_{j_2}$, $\beta_{j_1}$ and $\beta_{j_2}$ (with opposite signs if $\widehat{\boldsymbol{R}}_{j_1,j_2} < 0$), as well as $\psi_{j_1}$ and $\psi_{j_2}$ according to the following acceptance probability,

$$\begin{aligned} &\exp\left\{\left[\sum_{k\neq j_1,j_2}\left(\widehat{\boldsymbol{W}}_{k,j_1} - \widehat{\boldsymbol{W}}_{k,j_2}\right)\beta_k - \widehat{s}_{j_1}^{-2}\widehat{\beta}_{j_1} + \widehat{s}_{j_2}^{-2}\widehat{\beta}_{j_2}\right](\beta_{j_1} - \beta_{j_2})\right\} \\ &\times \exp\left\{\frac{1}{2}\left(\widehat{s}_{j_1}^{-2} - \widehat{s}_{j_2}^{-2}\right)\left(\beta_{j_1}^2 - \beta_{j_2}^2\right)\right\} \times \exp\left\{(a_{j_1} - a_{j_2})(\tau_{j_2} - \tau_{j_1})\right\}, \end{aligned} \tag{18}$$

if $\widehat{\boldsymbol{R}}_{j_1,j_2} > 0$, or

$$\begin{aligned}&\exp\left\{\left[\sum_{k\neq j_1,j_2}\left(\widehat{\boldsymbol{W}}_{k,j_1}+\widehat{\boldsymbol{W}}_{k,j_2}\right)\beta_k-\widehat{s}_{j_1}^{-2}\widehat{\beta}_{j_1}-\widehat{s}_{j_2}^{-2}\widehat{\beta}_{j_2}\right]\left(\beta_{j_1}+\beta_{j_2}\right)\right\}\\ &\times\exp\left\{\frac{1}{2}\left(\widehat{s}_{j_1}^{-2}-\widehat{s}_{j_2}^{-2}\right)\left(\beta_{j_1}^2-\beta_{j_2}^2\right)\right\}\times\exp\left\{\left(a_{j_1}-a_{j_2}\right)\left(\tau_{j_2}-\tau_{j_1}\right)\right\},\end{aligned} \tag{19}$$

if $\widehat{\boldsymbol{R}}_{j_1,j_2} < 0$.

(f) Repeat Steps (b)-(e) until convergence, and compute $h^2 = c^2\boldsymbol{\beta}^\top\boldsymbol{R}\boldsymbol{\beta}$ for each sample.

#### 3 Identifying credible sets

To get CSs, the coverage $\gamma$ and the minimum purity of CS $\rho$ should be specified. By default, $\gamma = 0.95$ and $\rho = 0.5$. We used a greedy algorithm to search all CSs with coverage $\geq \gamma$ and purity $\geq \rho$, as describe in Algorithm 1.

**Algorithm 1** Identify credible sets with coverage $\geq \gamma$ and purity $\geq \rho$.

**Require:** posterior samples of $\boldsymbol{\beta}$, estimated LD matrix $\widehat{\boldsymbol{R}}$

**Require:** coverage $\gamma \in [0, 1]$, purity $\rho \in [0, 1]$.

compute CLs for all SNPs.

sort all SNPs by descending order of CL. $\mathrm{CL}_{j_1} \geq \mathrm{CL}_{j_2} \geq \mathrm{CL}_{j_M}$.

$\mathcal{S} \leftarrow \{j_1, \ldots, j_M\}$, $\mathcal{T} \leftarrow \{\}$

**while** $\mathcal{S} \neq \emptyset$ **do**

  $k \leftarrow \mathcal{S}[1]$, remove $k$ from $\mathcal{S}$

  **if** $\mathrm{CL}_k \geq \gamma$ **then**

    output $\{k\}$ as a CS

  **else**

    $\mathcal{C} \leftarrow \{k\}$

    $\mathcal{V} \leftarrow$ all $l \in \mathcal{S} \bigcup \mathcal{T}$ such that $\left|\widehat{\boldsymbol{R}}_{k,l}\right| \geq \rho$

    $findCS \leftarrow$ **false**

    **while** $\mathcal{V} \neq \emptyset$ and $\mathrm{CL}_{\mathcal{C}} + \sum_{l \in \mathcal{V}} \mathrm{CL}_l \geq \gamma$ **do**

      $l \leftarrow \arg\max_{l \in \mathcal{V}} \mathrm{CL}_{\mathcal{C} \bigcup \{l\}}$

      $\mathcal{C} \leftarrow \mathcal{C} \bigcup \{l\}$

      **if** $\mathrm{CL}_{\mathcal{C}} \geq \gamma$ **then**

        output $\mathcal{C}$ as a CS, $findCS \leftarrow$ **true**

        $\mathcal{S} \leftarrow \mathcal{S} \setminus \mathcal{C}$, $\mathcal{T} \leftarrow \mathcal{T} \setminus \mathcal{C}$, break

      **else**

        $\mathcal{V} \leftarrow$ all $l \in \mathcal{S} \bigcup \mathcal{T}$ such that $\left|\widehat{\boldsymbol{R}}_{k',l}\right| \geq \rho$ for all $k' \in \mathcal{C}$

      **end if**

    **end while**

    **if** $!findCS$ **then**

      $\mathcal{T} \leftarrow \mathcal{T} \bigcup k$

    **end if**

  **end if**

**end while**

### Supplemental Figures

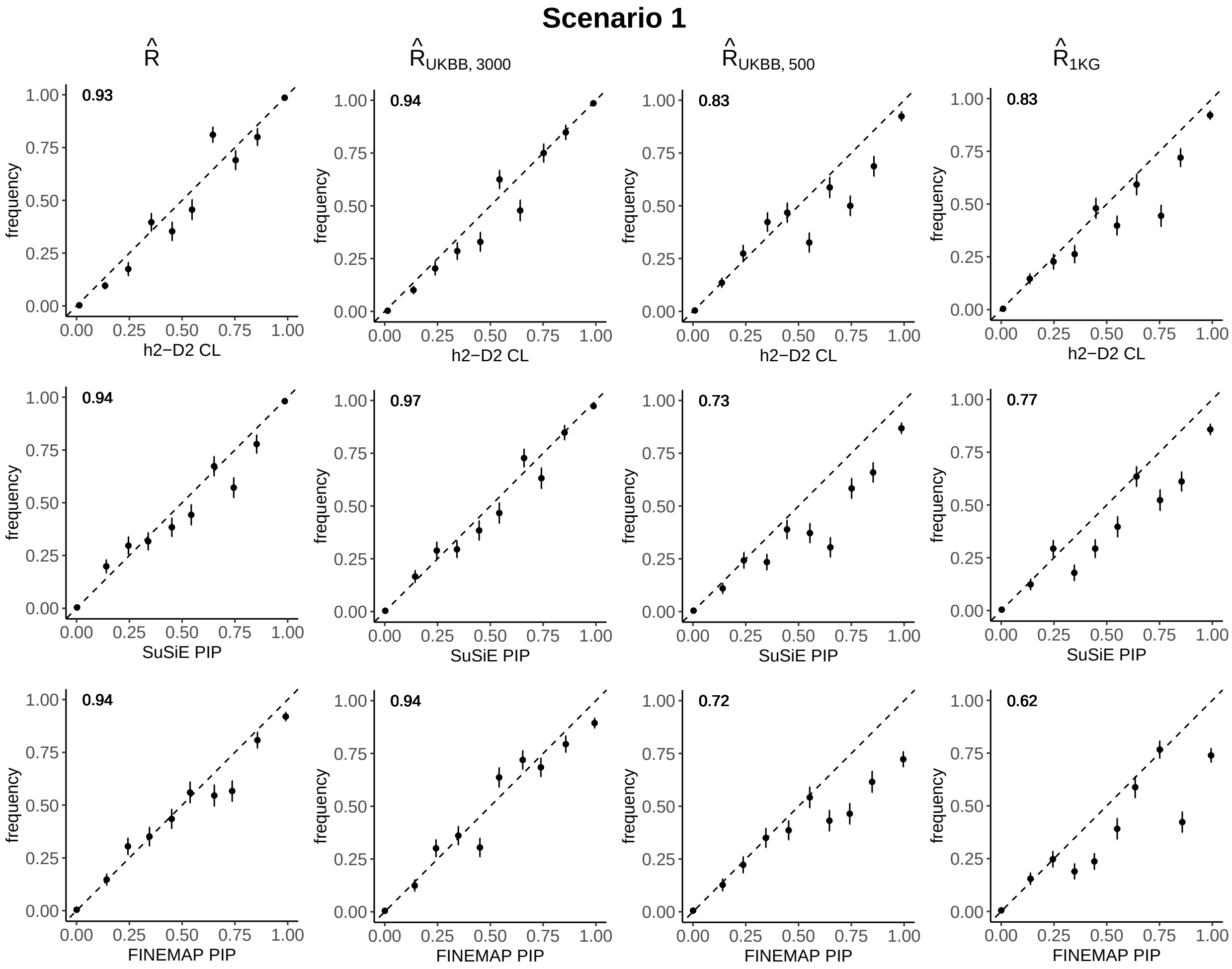

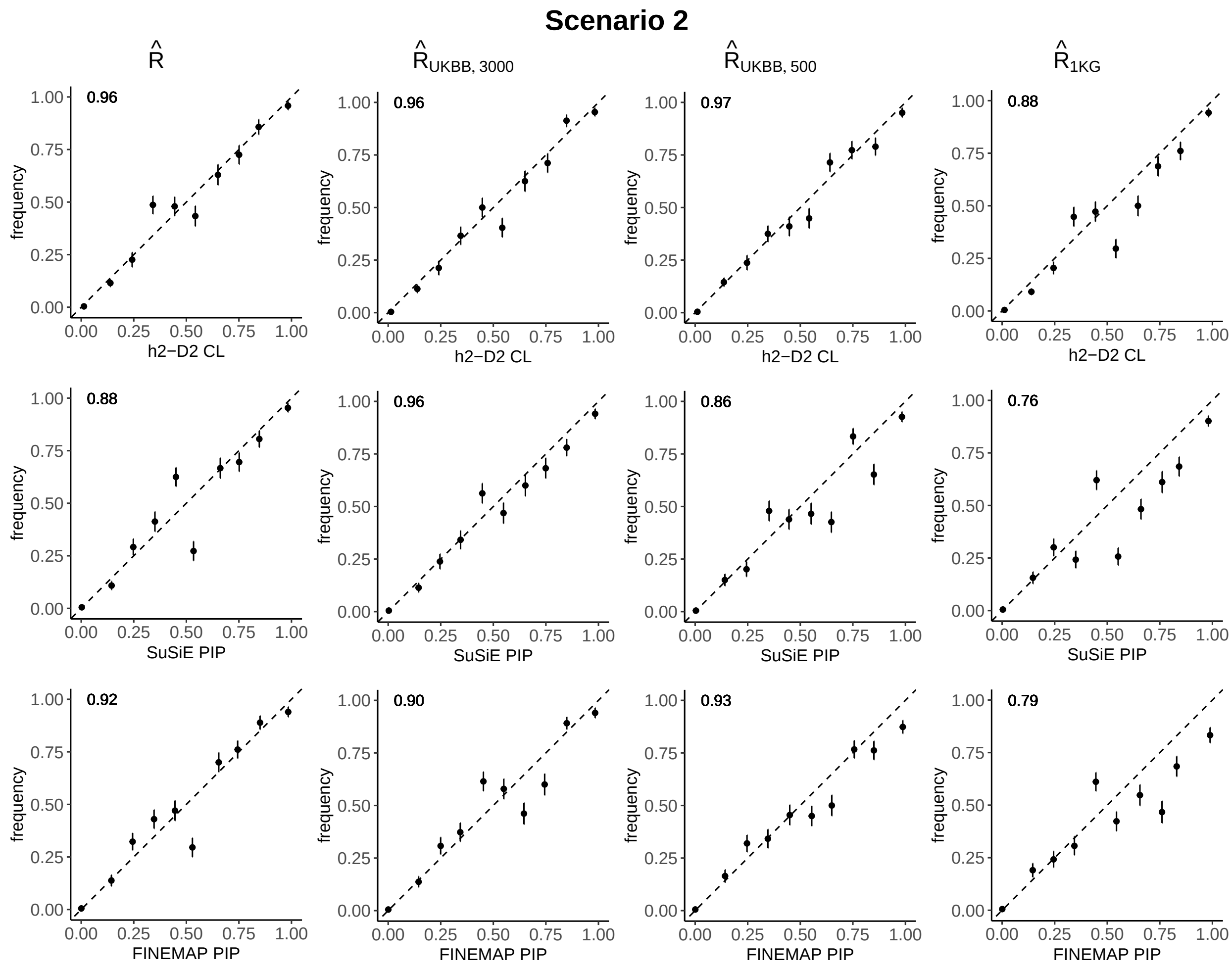
Scenario 2
$\hat{R}$
$\hat{R}_{UKBB,\,3000}$
$\hat{R}_{UKBB,\,500}$
$\hat{R}_{1KG}$
0.96
0.96
0.97
0.88
0.88
0.96
0.86
0.76
0.92
0.90
0.93
0.79
frequency
h2−D2 CL
SuSiE PIP
FINEMAP PIP
0.00
0.25
0.50
0.75
1.00

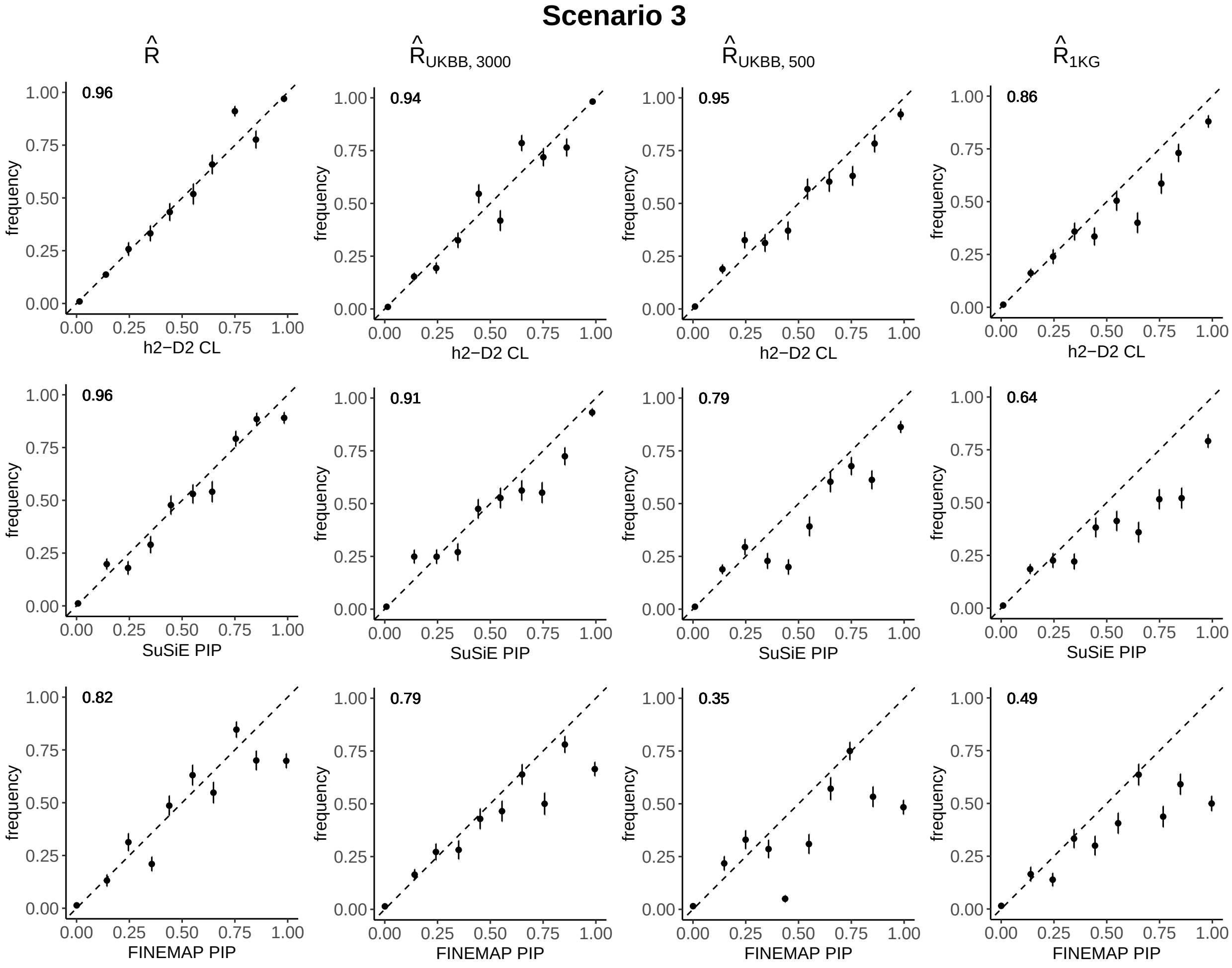
Scenario 3
$\hat{R}$
$\hat{R}_{UKBB,\,3000}$
$\hat{R}_{UKBB,\,500}$
$\hat{R}_{1KG}$
0.96
0.94
0.95
0.86
0.96
0.91
0.79
0.64
0.82
0.79
0.35
0.49
frequency
h2−D2 CL
SuSiE PIP
FINEMAP PIP
0.00
0.25
0.50
0.75
1.00

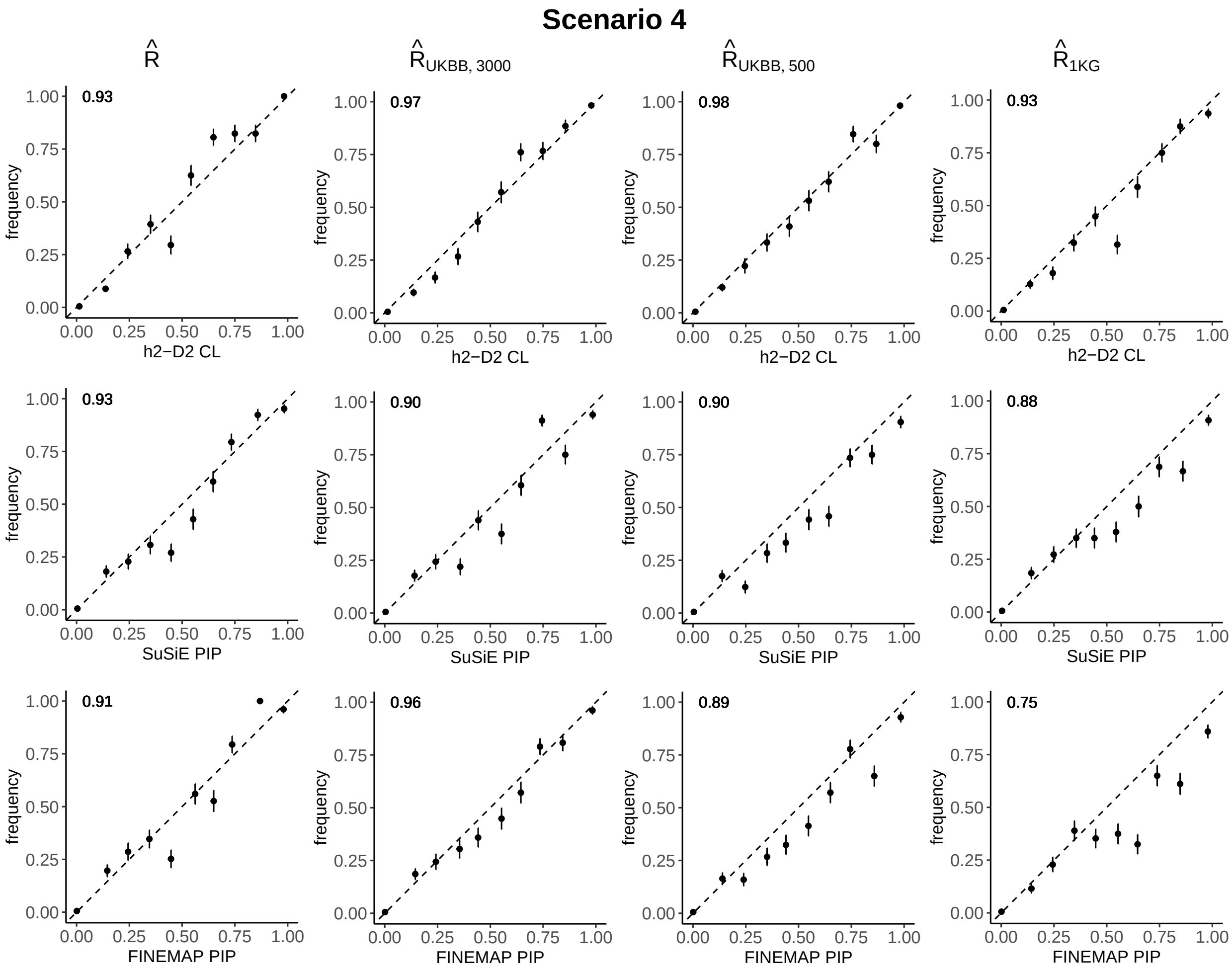


**Supplementary Fig. 1**: **Calibration of h2-D2 credible level, SuSiE PIP, and FINEMAP PIP.** For each simulated setting, SNPs were divided into 10 equally spaced bins from 0 to 1 based on their credible levels or PIPs. The plots illustrate the average proportion of causal variants across 100 simulated datasets against the average credible level or PIP within each bin. Error bars represent the standard errors across the 100 simulated datasets. $R^2$ is indicated in the upper left corner of each plot, providing a measure of the calibration quality.

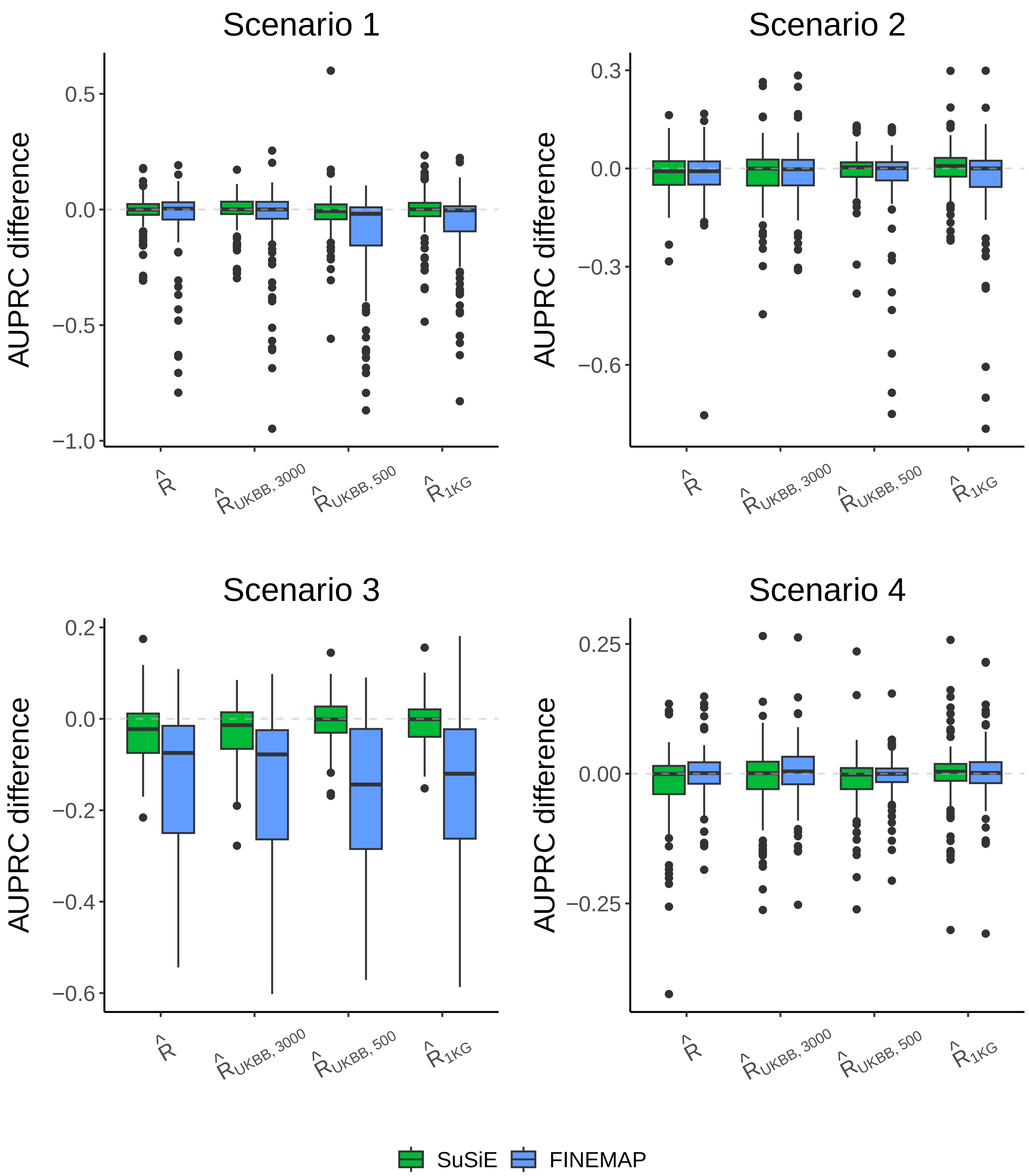


**Supplementary Fig. 2**: **Difference in AUPRC between h2-D2 and the other two methods across simulated datasets.** The boxplots demonstrate the variation in the difference of AUPRC values between h2-D2 and the other two methods (SuSiE and FINEMAP). Each point on the plot represents a simulated dataset. Green: AUPRC(SuSiE) - AUPRC(h2-D2). Blue: AUPRC(FINEMAP) - AUPRC(h2-D2).

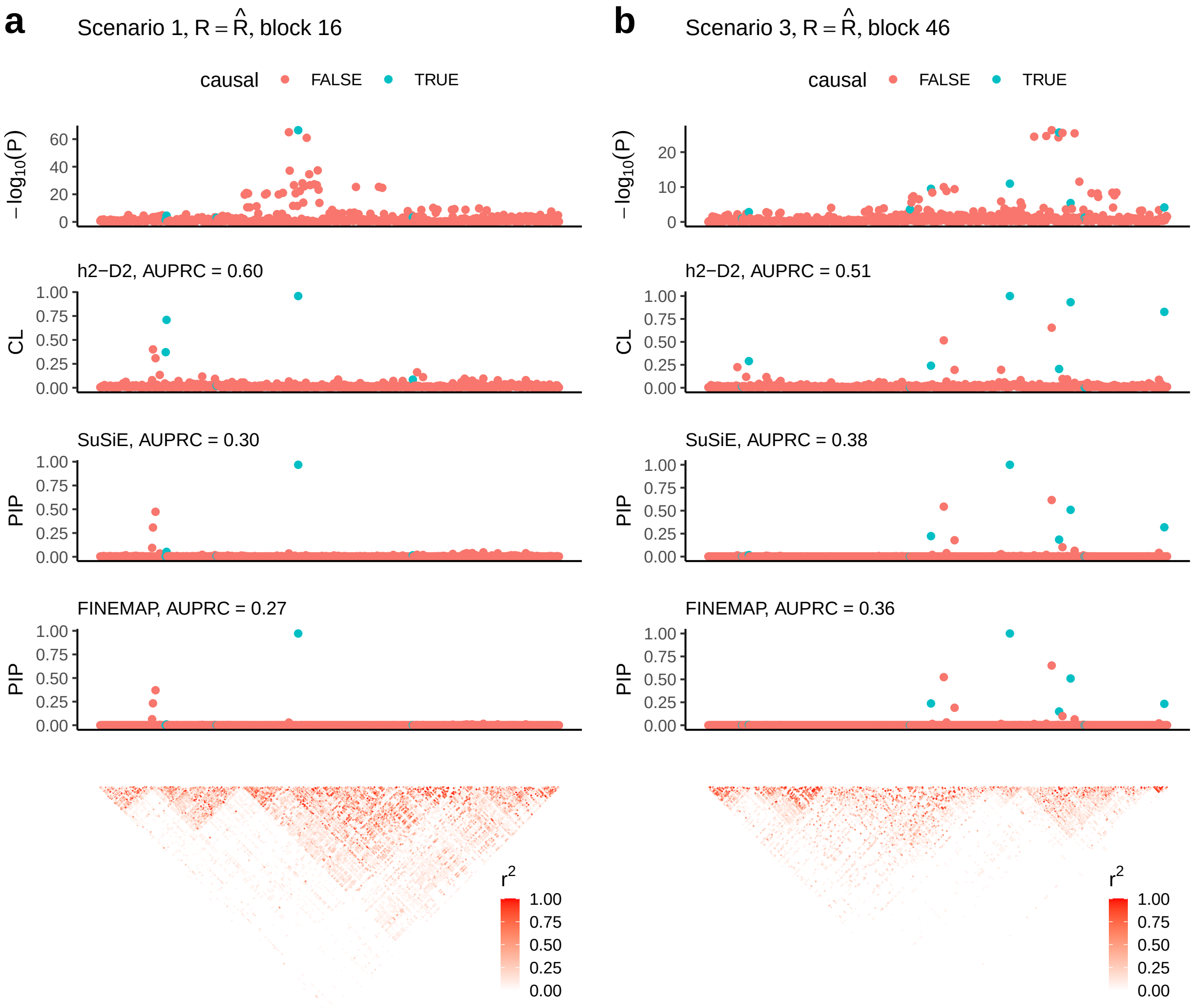


**Supplementary Fig. 3**: **Comparison of fine-mapping accuracy in two simulation datasets.** **a** Scenario 2, block 16 (chr2 18229255-18524162, $M = 541$). **b** Scenario 3, block 46 (chr2 105125906-106071159, $M = 681$). Each point represents a variant, with the blue points indicating causal variants. The first row displays the Manhattan plots indicating the marginal associations of variants ($-\log_{10}(P)$). The second row shows the credible levels of each SNP obtained from h2-D2. The third and fourth rows demonstrate the PIP of each SNP from SuSiE and FINEMAP, respectively. The bottom section depicts the LD heatmaps of the regions.

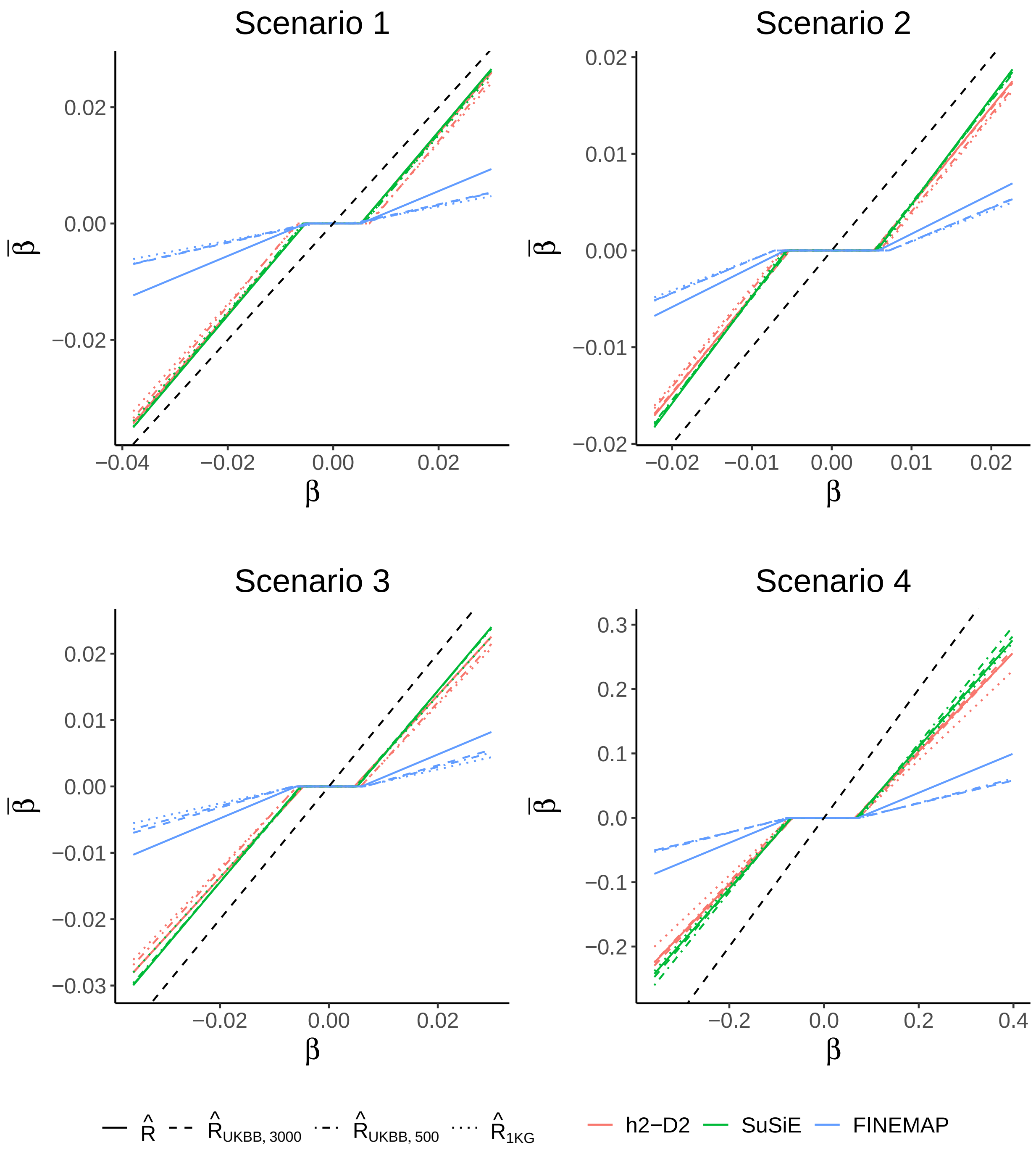


**Supplementary Fig. 4**: **Comparison of true values and posterior means of causal variant effect sizes.** The plot shows the relationship between the true values and posterior means of causal variant effect sizes, fitted by a piecewise linear model using all causal variants of 100 simulated datasets (Methods).

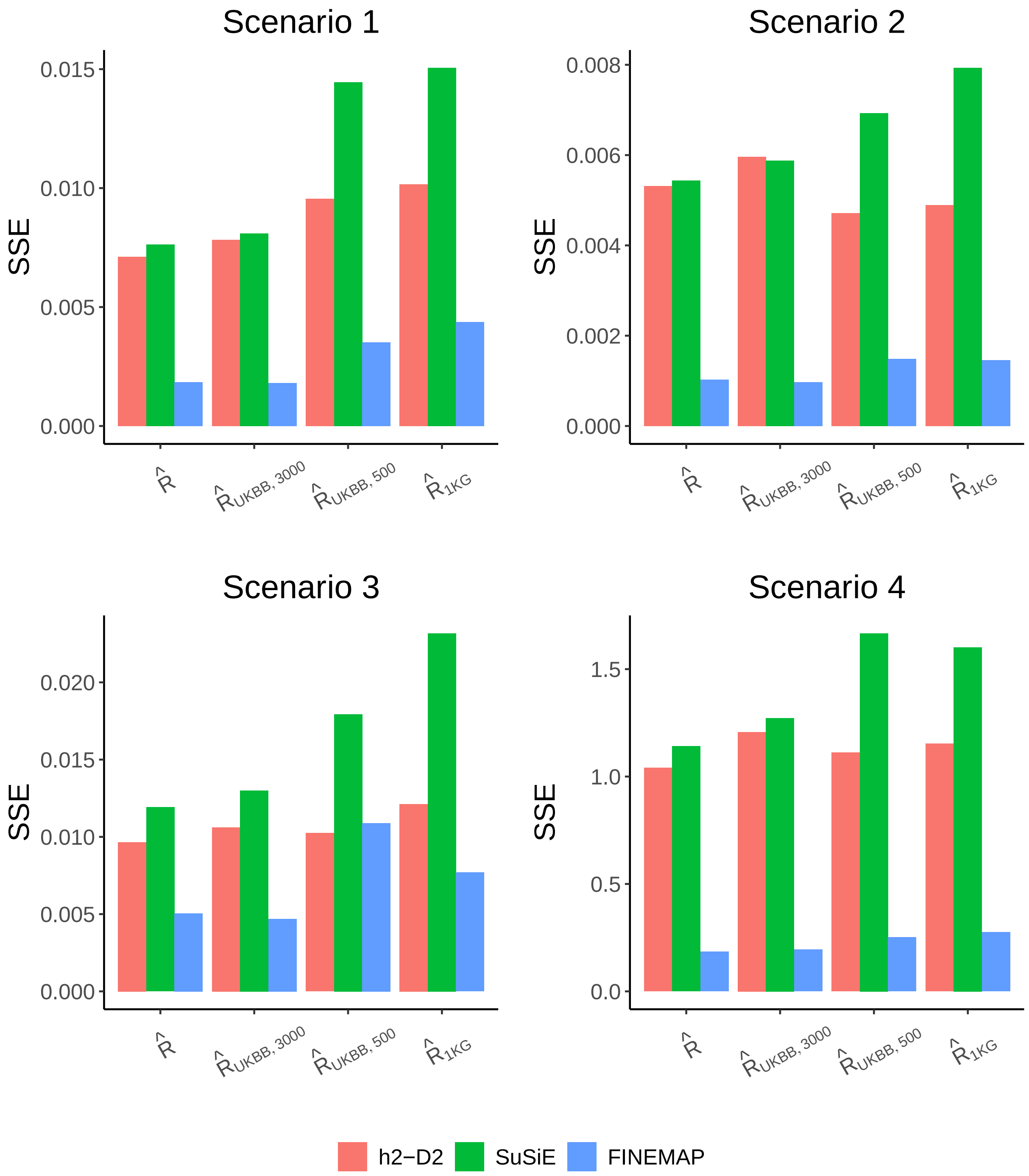


**Supplementary Fig. 5**: **Sum of squared error of effect sizes for non-causal variants based on posterior means.** The values represent the sum of squared errors across 100 simulated datasets for each setting.

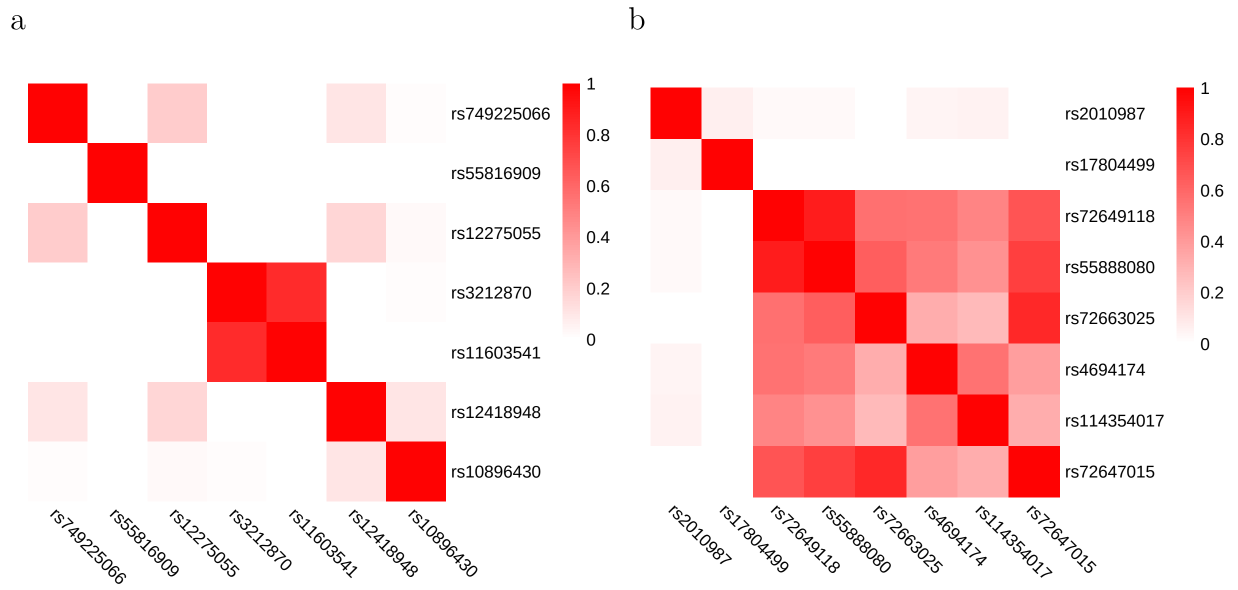


**Supplementary Fig. 6**: **LD heatmaps of tag SNPs in 95% CSs. a** LD heatmap of 5 tag SNPs from 4 CSs identified in chr11 68810837-69542062, along with two SNPs with credible levels $\geq 0.4$. **b** LD heatmap of 8 tag SNPs from 3 CSs identified in chr4 73256856-74885359.

### Supplementary Table

**Supplementary Table 1** (xlsx.): 100 blocks on chr2 selected for simulation studies.

**Supplementary Table 2** (csv.): Full list of variants in 95% credible sets identified by h2-D2 in prostate cancer fine-mapping analysis, as well as detailed annotations.

**Supplementary Table 3** (xlsx.): Enrichment of variants in 95% credible sets within DNaseI hypersensitive sites, ChIP-seq peaks of histone marks and transcription factor binding sites.

**Supplementary Table 4** (xlsx.): Estimated regression coefficients of linear model fitting $\log\left(\overline{\beta^2}\right)$ of tag SNPs by all functional annotations.

**Supplementary Table 5** (xlsx.): Enrichment of putative target genes regulated by credible causal variants within pathways from GO Biological Process and WikiPathways. Redundant pathways were removed (Methods).
